# Identification of transdiagnostic psychiatric disorder subtypes using unsupervised learning

**DOI:** 10.1101/2021.02.04.21251083

**Authors:** Helena Pelin, Marcus Ising, Frederike Stein, Susanne Meinert, Tina Meller, Katharina Brosch, Nils R. Winter, Axel Krug, Ramona Leenings, Hannah Lemke, Igor Nenadic, Stefanie Heilmann-Heimbach, Andreas J. Forstner, Markus M. Nöthen, Nils Opel, Jonathan Repple, Julia Pfarr, Kai Ringwald, Simon Schmitt, Katharina Thiel, Lena Waltemate, Alexandra Winter, Fabian Streit, Stephanie Witt, Marcella Rietschel, Udo Dannlowski, Tilo Kircher, Tim Hahn, Bertram Müller-Myhsok, Till F. M. Andlauer

**Author notes:** **Corresponding authors:** Dr. Till F. M. Andlauer, Klinikum rechts der Isar, Neuro-Kopf-Zentrum, Ismaninger Str. 22, 81675 Munich, Germany, E-Mail, Helena Pelin, Max Planck Institute of Psychiatry, Kraepelinstr. 2-10, 80804 Munich, Germany, E-Mail.

## Abstract

Psychiatric disorders show heterogeneous clinical manifestations and disease trajectories, with current classification systems not accurately reflecting their molecular etiology. This heterogeneity impedes timely and targeted treatment. Our study aimed to identify diagnostically mixed psychiatric patient clusters that share clinical and genetic features and may profit from similar therapeutic interventions. We used unsupervised high-dimensional data clustering on deep clinical data to identify transdiagnostic groups in a discovery sample (N=1250) of healthy controls and patients diagnosed with depression, bipolar disorder, schizophrenia, schizoaffective disorder, and other psychiatric disorders. We observed five diagnostically mixed clusters and ordered them based on severity. The least impaired cluster 0, containing most healthy controls, was characterized by general well-being. Clusters 1-3 differed predominantly regarding levels of maltreatment, depression, daily functioning, and parental bonding. Cluster 4 contained most patients diagnosed with psychotic disorders and exhibited the highest severity in many dimensions, including medication load. MDD patients were present in all clusters, indicating that we captured different disease stages or subtypes. We replicated all but the smallest cluster 1 in an independent sample (N=622). Next, we analyzed genetic differences between clusters using polygenic scores (PGS) and the psychiatric family history. These genetic variables differed mainly between clusters 0 and 4 (prediction AUC=81%; significant PGS: cross-disorder psychiatric risk, schizophrenia, and educational attainment). Our results confirm that psychiatric disorders consist of heterogeneous subtypes sharing molecular factors and symptoms. The identification of transdiagnostic clusters advances our understanding of the heterogeneity of psychiatric disorders and may support the development of personalized treatment regimes.

## Introduction

In the absence of biomarkers, common psychiatric disorders are typically diagnosed using reports and observations of an array of clinical symptoms. These disorders are multifactorial and constitute syndromic concepts with largely overlapping symptoms, resulting in heterogeneous clinical manifestations and disease trajectories. An individual’s genetic background shapes the susceptibility to psychiatric disorders to a large degree. Because all common, multifactorial disorders are highly polygenic, this risk can be quantified using polygenic scores (PGSs) [1]. Proportionally to the genetic risk load, a gradient of symptom severity may exist between healthy individuals and clinically diagnosed patients [2, 3].

The wealth of available data and advances in machine learning intensified efforts to redefine disorder categories using data-driven methods. Previous studies stratified psychiatric disorders mostly by clustering single domains (e.g., psychometry [4–8], neuroimaging [9–14], biochemical markers [15], or genetics [16, 17]) or disorders (e.g., major depressive disorder (MDD) [5, 9, 16, 18–20] or schizophrenia (SCZ) [21–26]). However, these analyses could not unravel shared neurobiological mechanisms, overlapping genetic profiles, and a transdiagnostic symptom continuum [27, 28].

Transdiagnostic clustering supports the existence of diagnostically mixed subtypes across two [4, 29–31] or more disorders [32–35]. However, previous transdiagnostic clustering studies were limited by small samples and analyzed few disorders or variables [36, 37]. To assess the continuum between well-being and disease, clustering analyses profit from the inclusion of healthy controls, largely omitted in previous studies [5, 19, 34].

In the present study, we applied a data-driven clustering approach to a transdiagnostic patient/control sample. It encompassed healthy controls and patients diagnosed with MDD, bipolar disorder (BD), schizophrenia, schizoaffective disorder (SZA), or other psychiatric disorders (see below). Our study had two main aims: First, to use high-dimensional data clustering (HDDC) [38] to identify stable transdiagnostic clusters. Here, we used psychopathology measures, personality traits, cognitive functioning, social functioning, attachment style, environmental exposures in childhood and youth, parental factors, and quality of life measures. Second, to characterize differences of clinical and genetic variables between clusters using supervised machine learning. Moreover, we analyzed the information gain of PGS compared to the family history of psychiatric disorders and replicated our clustering solution in an independent sample.

## Materials and Methods

### Sample description

FOR2107 is an ongoing multi-center study recruiting patients via in- and outpatient services in Marburg and Münster, Germany; healthy subjects were recruited via newspaper advertisements [39]. The study protocols were approved by the ethics committees of the Medical Schools of the Universities of Marburg and Münster, following the Declaration of Helsinki, and all participants provided written informed consent. All subjects underwent a structured clinical interview for DSM-IV axis I disorders (SCID-I) [40], administered by trained clinical raters.

All individuals recruited in the first phase of the study were eligible for the discovery sample (N=1623), N=855 independent individuals recruited subsequently were included in the replication. First, participants who had withdrawn their consent, with missing diagnosis, and relatives were excluded. Second, individuals with missing information in any of the variables used for clustering were excluded (Suppl Methods S1). Final sample sizes were N=1250 (discovery) and N=622 (replication). Age and diagnosis distributions differed between both samples (*p*=0.01, *p*=0.002, respectively), sex did not (*p=*0.16). Among diagnostic groups, the proportions of healthy controls (*p*=0.005) and MDD patients (*p*=0.003) differed significantly (Table 1 and Suppl Table S1).

**Table 1:** Characterization of the discovery sample and clusters.

| Variable | Discovery | Cluster 0 | Cluster 1 | Cluster 2 | Cluster 3 | Cluster 4 |
| --- | --- | --- | --- | --- | --- | --- |
| N | 1250 | 535 | 38 | 266 | 215 | 196 |
| **Demographics** |  |  |  |  |  |  |
| Age, mean (SD) | 35.1 (13.0) | 31.7 (11.9) | 38.6 (13.9) | 35.3 (12.7) | 37.9 (13.7) | 40.3 (12.8) |
| Gender, N (%) male | 483 (39%) | 205 (38%) | 9 (24%) | 106 (40%) | 74 (34%) | 89 (45%) |
| Years of education, mean (SD) | 13.5 (2.6) | 14.4 (2.4) | 13.3 (2.6) | 13.8 (2.6) | 13.1 (2.8) | 12.1 (2.7) |
| Living with partner, N (%) | 277 (28%) | 108 (20%) | 7 (18%) | 51 (19%) | 62 (29%) | 49 (25%) |
| BMI, mean (SD) | 25.3 (5.5) | 23.7 (4.3) | 24.9 (5.1) | 24.8 (4.9) | 27.0 (6.6) | 28.1 (6.4) |
| Family history (any psychiatric disorder), N (%) | 533 (43%) | 141 (26%) | 16 (42%) | 148 (56%) | 105 (49%) | 123 (63%) |
| **Diagnosis** |  |  |  |  |  |  |
| AAO*, mean (SD) | 25.2 (11.9) | 24.5 (9.9) | 29.6 (13.3) | 23.3 (11.5) | 27.8 (12.8) | 24.4 (11.4) |
| HC, N (%) | 590 (47%) | 448 (84%) | 19 (50%) | 78 (29%) | 34 (16%) | 11 (6%) |
| BD, N (%) | 75 (6%) | 9 (2%) | 1 (3%) | 15 (6%) | 26 (12%) | 24 (12%) |
| MDD, N (%) | 477 (38%) | 56 (10%) | 17 (45%) | 152 (57%) | 147 (68%) | 105 (54%) |
| SCZ, N (%) | 53 (4%) | 4 (1%) | 1 (3%) | 8 (3%) | 3 (1%) | 37 (19%) |
| SZA, N (%) | 25 (2%) | 2 (0.3%) | 0 (0%) | 5 (2%) | 2 (1%) | 16 (8%) |
| Other, N (%) | 30 (2%) | 16 (3%) | 0 (0%) | 8 (3%) | 3 (1%) | 3 (2%) |
| **Quality of life (SF36), mean (SD)** |  |  |  |  |  |  |
| General health | 66 (23.4) | 81.2 (14.0) | 57.9 (26.6) | 64.7 (19.8) | 49.4 (20.7) | 45.8 (21.3) |
| Mental health | 64.3 (22.6) | 81.4 (9.8) | 55.6 (23) | 59.4 (18.9) | 46.4 (19.3) | 45.6 (21.3) |
| **Depression and anxiety, mean (SD)** |  |  |  |  |  |  |
| HAMA sum | 7.3 (7.9) | 2.2 (2.5) | 9.7 (8.6) | 7.6 (6.4) | 13.1 (8.6) | 14 (8.8) |
| HAMD sum | 5.4 (6.6) | 1.1 (1.6) | 6.3 (6.7) | 5.9 (5.7) | 9.9 (7.0) | 11 (7.5) |
| BDI sum | 10.7 (10.8) | 3.2 (3.3) | 15.1 (12.8) | 12.7 (9.6) | 17.6 (10.1) | 20.3 (11.7) |
| **Positive, negative, and manic symptoms, mean (SD)** |  |  |  |  |  |  |
| SANS | 5.7 (9.9) | 0.6 (1.7) | 5.2 (8.6) | 6.9 (9.6) | 8.0 (9.1) | 15.6 (14.4) |
| SAPS | 1.4 (5.2) | 0.1 (0.6) | 0.1 (0.4) | 0.6 (1.6) | 0.7 (1.8) | 6.7 (11.5) |
| YMRS | 1.2 (2.5) | 0.5 (1.1) | 0.8 (1.4) | 1.1 (1.8) | 1.3 (2) | 2.9 (4.9) |
| **Maltreatment in Childhood and Youth (CTQ), mean (SD)** |  |  |  |  |  |  |
| Emotional abuse | 9.1 (4.7) | 6.3 (1.7) | 9.9 (4.9) | 11.4 (4.2) | 8.0 (3.0) | 14.6 (5.9) |
| Emotional neglect | 10.7 (5.2) | 7.5 (2.6) | 11.8 (5.4) | 14.1 (4.1) | 9.2 (3.5) | 16.5 (5.4) |
| Physical abuse | 6.2 (2.6) | 5.3 (0.7) | 6.4 (2.2) | 6.4 (2) | 5.5 (1.1) | 9.5 (4.6) |
| Physical neglect | 7.2 (2.7) | 5.8 (1.4) | 7.2 (2.2) | 8.0 (2.2) | 6.4 (1.6) | 10.4 (3.7) |
| Sexual abuse | 5.8 (2.5) | 5.1 (0.4) | 5.8 (2) | 5.8 (1.9) | 5.6 (1.7) | 8.0 (4.9) |

**Abbreviations:** BMI, body mass index; AAO, age at onset of any psychiatric disorder (defined according to OPCRIT item 4), \*not available for healthy controls; HC, healthy controls; BD, bipolar disorder; MDD,
major depressive disorder; SCZ, schizophrenia; SZA, schizoaffective disorder; Other, other psychiatric diagnoses including anxiety, adjustment, and substance use disorders; HAMA, Hamilton Depression Rating Scale; HAMD, Hamilton Anxiety Rating Scale; BDI, Beck Depression Inventory (BDI-II, self-reported), SANS/SAPS, Scale for the Assessment of Negative/Positive Symptoms; SF36; 36-Item Short Form Survey.
**Variable ranges and interpretations:** Quality of life [0-100, 0=disability]; HAMD 21 item [0-66, 0=not present]; HAMA [0-56, 0=not present]; BDI [0-63, 0=not present]; SAPS/SANS [0-86/0-80, 0=not present]; CTQ [5-25, 5=less severe experiences].

### Variables used for clustering and cluster description

Fifty-seven baseline variables were used for clustering and the description of clusters. Following a suggestion by Maj (2018) [36], we combined the assessment of symptoms and disease development at the current stage with variables capturing antecedent events, such as parental factors and early environmental factors, and concomitant variables such as cognitive functioning, social functioning (resilience), and personality traits. To avoid over-representation of some diagnostic aspects, we included only the main scores of clinical rating scales and psychometric questionnaires (Suppl Table S2). Eighteen additional variables were used for post-hoc characterization of clusters, including Global Assessment of Functioning (GAF) scores, medication, smoking, and sociodemographic variables (Fig. 1, Suppl Tables S1 and S5).

**Figure 1:**
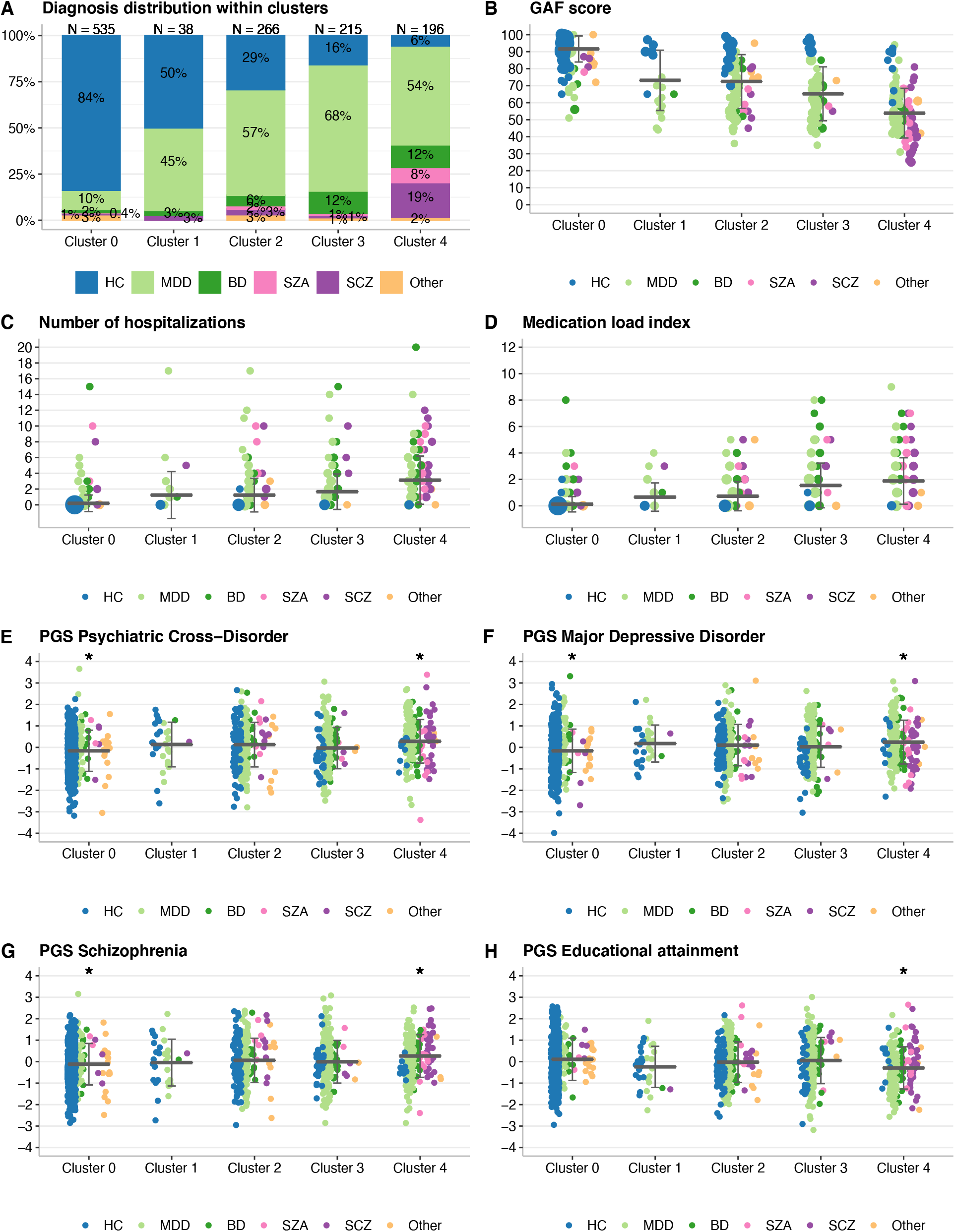
Cluster characterization in the discovery sample with clinical and genetic variables not used in the clustering pipeline. None of the variables shown in this Figure 1 or Supplementary Table S5 were included in the clustering pipeline. In panels B-H, a horizontal line represents the mean and the error bars indicate the standard deviation. The dot size is proportional to the number of individuals with the given value. Panels E-H show all PGS significant after Bonferroni correction (adjusted p<0.05), tested using the Westfall and Young procedure (Supplementary Methods S6), in either one-vs-all or one-vs-one analyses (Supplementary Tables S9-S10). PGS were standardized by Z-score transformation, the y-axis unit is standard deviations. **A:** The distribution of diagnoses within clusters. **B:** The Global Assessment of functioning (GAF) score, used for sorting clusters. Lower scores imply more severe impairment. **C:** The number of times an individual was hospitalized. **D:** The medication load index [59], reflecting the dose and variety of different medications taken. **E:** Psychiatric cross-disorder PGS, significantly different in two one-vs-all analyses (lower in Cluster 0, Bonferroni-corrected p=0.004; higher in Cluster 4, corrected p=0.01). **F:** MDD PGS, significantly different in two one-vs-all analyses (lower in Cluster 0, p=0.008; higher in Cluster 4, corrected p=0.04). **G:** Schizophrenia PGS, significantly different in two one-vs-all analyses (lower in Cluster 0, corrected p=0.04; higher in Cluster 4, corrected p=0.01). **H:** Educational attainment PGS, significantly different in one one-vs-all analysis (lower in Cluster 4, corrected p=0.004).

The self-reported family history of either any psychiatric disorder or specifically for MDD, BD, and SZA/SCZ was assessed for first-degree relatives and used for the genetic cluster characterization. We contrasted known with no/unknown family history.

### Genotyping and calculation of PGS

Genotyping was conducted using the PsychArray BeadChip, followed by quality control and imputation, as described previously [41, 42] (Suppl Methods S2). Imputed genetic data were available for n=1146 discovery-stage and n=556 replication-stage individuals (Suppl Fig. S1). PGSs were calculated for ten disorders and traits using PRS-CS [43] (Suppl Methods S3) with training data from sufficiently powered, published genome-wide association studies (GWASs): attention-deficit/hyperactivity disorder (ADHD) [44], autism spectrum disorder (ASD) [45], BD [46], psychiatric cross-disorder (CD) [47], educational attainment (EA) [48], extraversion [49], hedonic well-being (HWB) [50], MDD [51], neuroticism [52], and schizophrenia [53].

### Clustering analysis

The clustering of discovery-stage scaled clinical variables was conducted by HDDC [38] using the *R* (v3.6.0) package *HDclassif* [54]. This package implements a subspace clustering algorithm based on the Gaussian mixture model framework, which allowed us to fit 14 different model types, corresponding to different regularizations for the cluster solutions. The clustering pipeline had four steps: finding the best fitting model type, finding the optimal cluster number, getting the final cluster solution, and assessing the solution’s stability (Suppl Methods S4 and Suppl Fig. S2).

### Characterization of clusters

In primary analyses, we characterized the clusters with the *one-vs-all* strategy [55], with *one-vs-one* pairwise comparisons in secondary analyses. Genetic analyses used 24 variables: 10 PGS, 4 family history, 8 ancestry components, age, and gender. Merged with family history, the genetic sample size was n=1137 (discovery) and n=542 (replication). We analyzed by supervised high-dimensional discriminant analysis (HDDA) [54, 56] which of the 57 variables used for clustering were most important for the cluster characterization. Lasso regularized regression [57] was used to predict cluster labels with genetic variables (Suppl Methods S5). Statistical testing was performed using the Westfall and Young method [58], controlling the family-wise error rate while accounting for the possible dependence structure of the analyzed variables. The obtained *p*-values were subsequently corrected for the number of comparisons using Bonferroni’s method. For thus adjusted *p*-values, a significance threshold α=0.05 was used (Suppl Methods S6). We used multinomial regression to compare PGS with family history when predicting clusters (Suppl Methods S7).

### Replication analysis

We clustered the replication sample using the discovery-stage model parameters (Suppl Methods S8). Discovery-stage *one-vs-all* HDDA classification models were fit to the replication-stage clusters. Replication clusters were identified using the best discovery-stage model (balanced accuracy >70%). Genetic prediction analyses were conducted using discovery-stage *one-vs-all* lasso regression models.

## Results

### Model-based clustering analysis

The discovery-stage dataset contained N=1250 individuals with a mean age of 35.1 (SD=13.0) years. For the distribution of diagnoses, see Table 1. We performed model-based HDDC clustering using 57 baseline variables (Suppl Table S2). Our clustering pipeline (Suppl Fig. S3) identified five clusters (Fig. 1A), which were ordered by their average GAF scores, from lowest (cluster 0) to highest severity (cluster 4) (Fig. 1B).

### Phenotypic characterization of clusters

Cluster 0 contained mostly healthy controls, while the other clusters were diagnostically more mixed (Fig. 1A). All clusters showed distinct profiles of diagnoses, symptoms, and environmental risk factors (Table 1, Suppl Table S3).

Individuals in cluster 0 (n=535, 84% healthy controls) showed the overall best health and quality of life and exhibited the lowest severity in most symptom and risk scores (Figs. 1-2, Suppl Table S4-S5). The smallest cluster 1 (n=38) included the highest rates of females (62%) and symptomatic controls without a diagnosis (50%), who reported reduced general and mental health and increased anxiety and depression symptoms (Table 1, Fig. 2). Individuals in cluster 2 showed average general health scores but reduced mental health and parental bonding and elevated emotional maltreatment scores (Table 1, Fig. 2, Suppl Table S4). Cluster 3 had the highest rate of affective diagnoses with high depression and anxiety levels (Fig. 1A, Table 1); its members reported substantially reduced general and mental health. Childhood maltreatment scores in cluster 3 did not differ from the overall average (Table 1). Cluster 4 (n=196) featured most patients diagnosed with SZA and schizophrenia (Fig. 1A). Individuals in cluster 4 were characterized by the highest severity in many dimensions used for clustering (Table 1, Fig. 2, Suppl Table S4) and in additional variables examined post-hoc, such as hospitalization and medication load index [59] (Fig. 1, Suppl Table S5).

**Figure 2:**
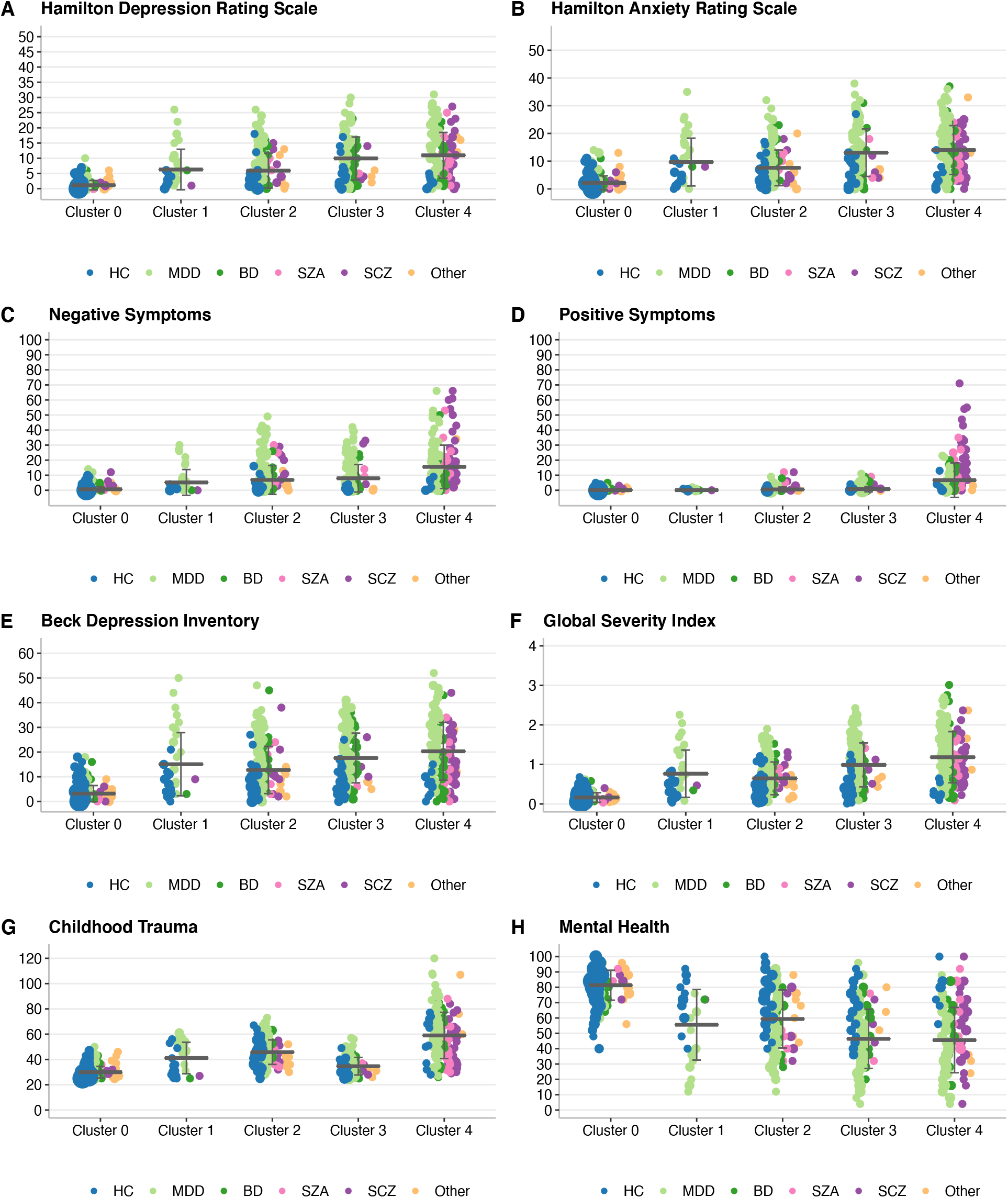
Cluster characterization in the discovery sample with variables used in the clustering pipeline. A horizontal line represents the mean, and the error bars indicate the standard deviation, while the dot size is proportional to the number of individuals with the given value. **A:** Hamilton Depression Rating Scale (HAMD, 21 items, clinician-administered), range 0-66, scores >7 indicate (mild) depression. **B:** Hamilton Anxiety Rating Scale (HAMA), range 0-56, scores >17 indicate mild to moderate anxiety severity. **C:** Scale for the Assessment of Negative Symptoms (SANS, sum score), range 0-80, a higher score indicates more severe negative symptoms. For subscales, see Suppl Table S5. **D:** Scale for the Assessment of Positive Symptoms (SAPS, sum score), range 0-86, a higher score indicates more severe positive symptoms. For subscales, see Suppl Table S5. **E:** Beck Depression Inventory (BDI-II, self-reported), range 0-63, scores >9 indicate (mild) depression. **F:** Symptom Checklist – Global Severity Index, an index of overall psychological distress, range 0-4, higher scores reflect higher levels of psychopathological distress as well as a greater severity of self-reported symptoms. **G:** Childhood Trauma Questionnaire sum score, range 25-125, a higher score indicates more experiences of childhood trauma. **H:** SF36 – Quality of life measurements – Mental health, range 0-100, high scores define a more favorable health state.

As a secondary analysis, we characterized MDD patients within the five clusters to assess the heterogeneity of this large diagnostic group and identified distinct phenotypic signatures of MDD patients in each cluster (Suppl Tables S6-S7).

### Genetic characterization of clusters

We conducted lasso regularized regression to predict cluster assignments using genetic variables, *i*.*e*., ten PGS and four self-reported family history assessments. Prediction performances were highest for the two extreme clusters 0 and 4 (*cluster 0 vs. 4*: *AUC=81%, sensitivity=75%, specificity=75%; cluster 0 vs. all*: *AUC=71%, sensitivity=66%, specificity=66%; cluster 4 vs. all*: AUC=73%, *sensitivity=67%, specificity=67%*, Suppl Table S8). Lasso selected seven variables when comparing cluster 0 against all others and 16 for cluster 4 (Table 2). In both cases, the self-reported family history achieved larger effect sizes than PGSs of psychiatric disorders. *One-vs-all* comparisons of clusters 0, 2, and 4 showed significant genetic variables (Table 2, Suppl Table S9 and Fig. 1 E-H): Cluster 0 was characterized by a lower family history of MDD, BD, and any psychiatric disorder (each adjusted *p*=0.004) and lower cross-disorder (*p*=0.004), MDD (*p*=0.008), and schizophrenia (*p* =0.04) PGS. Cluster 2 was characterized by a higher family history of any psychiatric disorder *p*=0.005) and MDD (*p*=0.03). Cluster 4 showed a higher family history of any psychiatric disorder (*p*=0.004) and higher cross-disorder (*p*=0.01), schizophrenia (*p*=0.01), and MDD (*p*=0.04) PGS, as well as lower PGS for educational attainment (*p*=0.004). Pairwise comparisons resulted in significant differences between four cluster pairs (Suppl Table S10). Cluster 4 MDD patients showed significantly higher ADHD (*p*=0.01) and lower educational attainment PGS (*p*=0.005) than MDD patients from the other clusters (Suppl Table S6 and Suppl Fig. S4 A-B).

**Table 2:** Genetic characterization of the discovery clusters.

| Category | Lasso model coefficients | Cluster 0 vs. all | Cluster 1 vs. all | Cluster 2 vs. all | Cluster 3 vs. all | Cluster 4 vs. all |
| --- | --- | --- | --- | --- | --- | --- |
| Demographic | Age | **-0.335** | 0.077 | 0 | 0.181 | **0.381** |
|  | Gender | 0 | -0.188 | 0 | -0.081 | 0.139 |
| Ancestry components | AC1 | 0 | 0 | 0 | 0.081 | 0 |
|  | AC2 | 0 | 0 | 0 | 0.013 | 0 |
|  | AC3 | 0 | 0.061 | 0 | 0.003 | -0.063 |
|  | AC4 | 0 | 0.122 | 0 | 0 | -0.144 |
|  | AC5 | 0 | 0.086 | 0 | -0.03 | 0.083 |
|  | AC6 | 0 | 0 | 0 | 0 | -0.004 |
|  | AC7 | 0 | 0 | 0 | -0.069 | 0 |
|  | AC8 | 0 | 0 | 0 | -0.003 | 0 |
| Family history | Any psychiatric disorder | **-0.445** | 0 | **0.159** | 0 | **0.338** |
|  | BD | **-0.007** | -0.06 | 0 | 0 | 0.238 |
|  | MDD | **0** | 0 | **0.044** | 0.095 | 0 |
|  | Schizophrenia | 0 | 0 | 0 | 0.029 | 0.051 |
| Polygenic scores | Cross psychiatric disorder | **-0.093** | 0.102 | 0 | -0.04 | **0.093** |
|  | ADHD | 0 | 0 | 0 | -0.018 | 0 |
|  | ASD | 0 | 0 | 0 | 0 | 0 |
|  | BD | 0 | -0.199 | 0 | 0.094 | 0 |
|  | MDD | **-0.074** | 0.05 | 0 | 0 | **0.112** |
|  | Schizophrenia | **0** | 0 | 0 | 0 | **0.167** |
|  | Educational attainment | 0.043 | -0.094 | 0 | 0.002 | **-0.269** |
|  | Extraversion | 0 | 0 | 0 | 0.023 | 0.107 |
|  | Hedonic wellbeing | 0 | 0 | 0 | -0.018 | -0.128 |
|  | Neuroticism | -0.009 | 0 | 0.05 | 0.095 | -0.016 |

**Abbreviations:** AC, ancestry components; BD, Bipolar Disorder; MDD, Major Depressive Disorder; ADHD, attention-deficit/hyperactivity disorder; ASD, Autism spectrum disorder.

### Assessment of the information gain by genetic variables

The inclusion of PGSs and ACs in a multinomial cluster prediction model yielded an increase of *R*^*2*^=11.7% over a null model without genetic variables (Suppl Table S11). The family history alone improved the *R*^*2*^ by 10.8% over the null model; a model with both family history and ACs showed a gain of *R*^*2*^=13.9%. PGSs, ACs, and family history together increased *R*^*2*^ by 20.3%. PGSs improved the model containing family history and ACs significantly (likelihood ratio test *p*=5×10^−5^).

### Replication of the clustering analysis

The replication dataset contained N=622 individuals with a mean age of 36.3 (SD=12.6) years (Suppl Table S1). HDDA models matched all but the smallest cluster 1 between discovery and replication samples (Suppl Fig. S5). The matched replication clusters followed the same severity ranking as the discovery-stage clusters, and many variables showed highly similar severity patterns (Fig. 3, Suppl Tables S1, S12, and S13).

**Figure 3:**
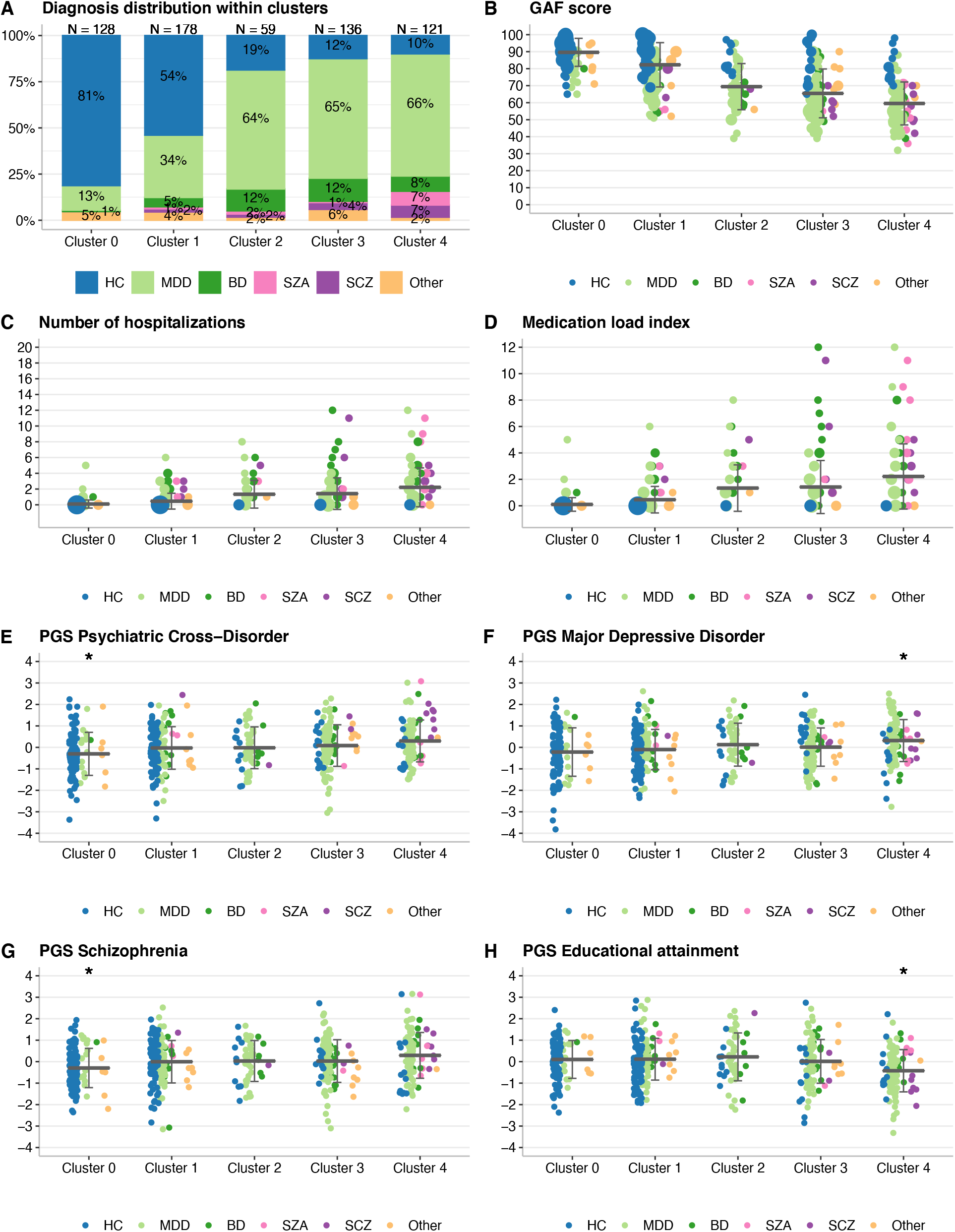
Cluster characterization in the replication sample with clinical and genetic variables not used in the clustering pipeline. In B-H, a horizontal line represents the mean, and the error bars indicate the standard deviation, while the dot size is proportional to the number of individuals with the given value. Panels E-H show all PGS that were significant after Bonferroni correction (adjusted *p*<0.05) in either *one-vs-all* or *one-vs-one* analyses using the Westfall and Young procedure (Supplementary Methods S6) in the discovery-stage analysis. All *p*-values for the full replication sample are shown in Suppl Tables S15 and S16. PGS were standardized by Z score transformation, the y-axis unit are standard deviations. **A**: The distribution of diagnoses within clusters. **B**: The Global Assessment of functioning (GAF) score, used for sorting clusters. Lower scores imply more severe impairment. **C**: The number of times an individual was hospitalized. **D**: Medication load index [59], reflecting dose and variety of different medications taken. **E**: Psychiatric cross-disorder PGS, replicated for the comparison *cluster 0-vs-all* (corrected *p*=0.03). **F**: Major depressive disorder PGS, replicated for the comparison *cluster 4-vs-all* (corrected *p*=0.01). **G**: Schizophrenia PGS, replicated for the comparison *cluster 0-vs-all* (*p*=0.005). **H**: Educational attainment PGS, replicated for the comparison *cluster 4-vs-all* (corrected *p*=0.005).

Projection of the genetic lasso regularized regression models showed an AUC=63%, sensitivity=60%, specificity=60% for cluster 0 *vs*. all and an AUC=68%, sensitivity=67%, specificity=66% for cluster 4 *vs*. all, a highly similar performance compared to the discovery sample. Further projections of five pairwise models yielded AUCs >60% (Suppl Table S14). As observed in the discovery sample, cross-disorder (adjusted *p*=0.03) and schizophrenia (*p*=0.005) PGS were significantly lower in the replication-stage cluster 0 (Suppl Table S15 and Fig. 3 E,G). For cluster 4, the MDD PGS (*p*=0.01) was higher and the educational attainment PGS (*p*=0.005) lower, confirming the discovery-stage results (Fig. 3 F,H). Also schizophrenia and crossdisorder PGS were, as in the discovery stage, higher in cluster 4, but these associations showed only nominal significance and did not pass correction for multiple testing. In pairwise comparisons, replicated PGS associations included the associations of schizophrenia, cross-disorder, and educational attainment PGS when comparing cluster 0 with 4 (Suppl Table S16). MDD individuals in cluster 4 had, as in the discovery stage, significantly lower EA PGS than MDD patients in other clusters, while the association of ADHD PGS for MDD patients in cluster 4 did not replicate (Suppl Fig. S4 C-D).

## Discussion

The symptoms and disease courses of patients diagnosed with any given major psychiatric disorder are highly heterogenous, suggesting ethiopathological differences between patients sharing the same diagnosis. The classification and treatment of psychiatric disorders rely on a nosological approach that does not necessarily reflect the disorders’ molecular etiology. In the present study, we applied a high-dimensional data clustering algorithm using deep phenotypic data on a transdiagnostic sample, including healthy controls, to characterize transdiagnostic psychiatric subgroups. We identified five diagnostically mixed clusters, which were ranked along a continuous severity scale. Cluster 0 contained mostly healthy controls and was distinguished by the lowest severity in many measures – from the lowest maltreatment factors, depression level, and positive symptoms to the highest quality of life scores. Cluster 4 had the highest share of schizophrenia and SZA patients and showed the highest severity in many variables not used for the clustering, e.g., the medication load index [59] and the number of hospitalizations. Clusters 1-3 ranged between these two extremes and differed mostly in different levels of maltreatment, depression and antidepressant use, daily functioning, and parental bonding.

Importantly, all but the smallest of these clusters were replicated in an independent sample. Given that the proportions of diagnoses in the replication sample differed, the replication of these clusters and their characteristics, especially the severity spectrum and genetic variables, is remarkable. It underlines the stability of the cluster solution and indicates that our approach did not suffer from overfitting in the discovery sample.

Supervised analyses of genetic variables confirmed that PGS added information to cluster comparisons beyond what could be assessed using the family history of disorders. The slight increase of explained variance conveyed by ancestry information underlined the highly polygenic nature of psychiatric disorders. Psychiatric cross-disorder, schizophrenia, and MDD PGS were significantly higher in the most severe cluster 4 compared to cluster 0, whereas educational attainment PGS were lower – corresponding to effect directions reported in previous studies [47, 51, 60–62].

Compared to DSM-IV diagnostic categories, our cluster solution surpassed diagnostic boundaries mostly for MDD and BD, while patients diagnosed with schizophrenia and SZA were primarily grouped in the high severity cluster 4. This finding confirms etiological similarities between the affective disorders MDD and BD, distinguishing them from predominantly psychotic disorders [62, 63]. MDD patients were present in all five clusters, suggesting that different disorder subtypes or stages were captured. Interestingly, 80% of MDD patients in the lowest severity cluster 0 were in remission of either single or recurrent MDD at the assessment time (coded according to the DSM). Hence, their present clinical presentation was similar to healthy individuals. MDD patients in cluster 1 might represent a reactive depression subtype, with similarities to burnout (*i*.*e*., a high somatization level and life stress, low energy, and a higher age of disorder onset). MDD cases in cluster 2, with the lowest average age of onset, might suffer from exogenous depression triggered by external stressors (maltreatment and neglect in childhood). Interestingly, this cluster also contained the highest ratio of BD type-II/type-I patients (Suppl Table S17). However, these patients also showed a high genetic predisposition for depression, with 48% reporting an MDD family history. In cluster 3, MDD patients showed a low influence of adverse environmental factors and high parental bonding, similar to cluster 0. Nevertheless, their quality of life was impacted negatively by illness – cluster 3 MDD patients showed low energy and experienced limitations in role activities because of physical and emotional health problems. Consistent with the strong presence of schizophrenia patients, cluster 4 MDD patients exhibited depression with psychotic features, showing higher positive symptoms and more antipsychotic intake. These MDD patients had significantly higher ADHD PGS than MDD patients in other clusters (*p*=0.009). Previous studies have identified correlations between ADHD in childhood and the development of other severe psychiatric disorders, especially schizophrenia, in adulthood [64–66]. Although not available at present, a retrospective assessment of ADHD symptoms during childhood in cluster 4 MDD cases might shed further light on this correlation.

Healthy controls distributed across clusters 1-4 showed isolated symptoms similar to the psychiatric patients in these clusters. The number of healthy controls decreased with cluster severity. Apparently, the symptoms of these healthy individuals were not sufficiently severe to generate a clinically relevant presentation of any psychiatric disorder fitting the currently used nosology. For example, these individuals may have only experienced short-term symptoms, e.g., resulting from a recent adverse life event. Indeed, healthy controls in cluster 4 showed a negative events score of 21, higher than the median of any other disorder group in the clusters showing high impairment. Alternatively, they might develop a disorder later in life; with a mean age of 32, the healthy individuals were younger than the average assessed patients. Follow-up assessments of the longitudinal FOR2107 study may reveal whether a higher share of healthy controls mapping to the more severe clusters will develop a disorder over time. Possibly, the current diagnostic criteria do not capture the whole illness spectrum. Our study might thus contribute to improved diagnostic criteria, as envisioned by the Research Domain Criteria (RDoC) project [67].

To our knowledge, the present study is the first to cluster multidomain profiles of clinical variables across psychiatric disorders and including healthy controls. Nevertheless, the cluster profiles and identified severity spectrum partially aligns with previous findings. A transdiagnostic study identified a cluster containing mainly healthy controls and exhibiting the lowest symptom scores in the observed dimensions [34], likely corresponding to our cluster 0. Our highly impaired cluster 4, with its high percentage of schizophrenic patients, low functioning, and significantly lower EA PGS, may correspond to the severe psychosis subtype from a previous study [32]. Moreover, a single-disorder subtyping study [5] detected five clusters of MDD exhibiting different symptom severities, with one subtype showing an absence of many symptoms, similar to our cluster 0. Furthermore, our results highlight the correlation of various measures of childhood trauma, adverse experiences, and lack of support with illness severity, positive symptoms, hospitalizations, and the need for more intensive treatment. Several prior studies support such a correlation [30, 68–71].

One limitation of our transdiagnostic clustering study was that most psychiatric patients were diagnosed with MDD, with only a smaller share of other, especially psychotic diagnoses. While the high number of MDD patients allowed for a detailed description of depression subtypes, a similar characterization was not possible for psychotic disorders, which concentrated in cluster 4. Other transdiagnostic studies applying our clustering approach with more psychotic patients could focus on BD and schizophrenia subtypes, as suggested by previous single-disorder studies [23, 26]. Moreover, although we used independent individuals for the replication dataset, these probands were subsequently recruited within the same study as the discovery-stage sample. Accordingly, the proportions of healthy controls and MDD patients differed between the discovery and replication samples, limiting their comparability. We conducted the quality control of the phenotypic and genetic data jointly for both datasets, introducing minor dependencies. Finally, the replication sample was smaller than the discovery sample, attenuating the statistical power. In conclusion, our study constitutes a data-driven, computational approach to psychiatric disorder stratification that surpasses existing diagnostic categories and integrates different domain profiles.

Our analyses support the hypothesis that psychiatric disorders consist of heterogeneous subtypes that share etiological factors and symptoms. We have demonstrated the importance of stratifying symptoms and disorder subtypes along a severity continuum. Individuals formally diagnosed with the same disorder differ in their specific impairment. Furthermore, their symptoms may partly overlap with symptoms exhibited by patients with different diagnoses, highlighting the need for symptomin-stead of diagnosis-specific treatment. Our transdiagnostic clustering approach may advance the understanding of the heterogeneity within and between psychiatric disorders. If applied to further cohorts, it may help the identification of patient groups sharing clinical features and thus profiting from similar treatments. The identification of such groups can lead to the development of more appropriate diagnoses, targeted treatment options, and prediction models for the disease course. Future assessments in FOR2107 and other longitudinal studies can reveal whether patients mapping to the different clusters show similar disease courses and treatment responses.

## Data Availability

Data is available from the corresponding authors on reasonable requests.

## Acknowledgments

This work is part of the German multi-center consortium “Neurobiology of Affective Disorders. A translational perspective on brain structure and function”, funded by the German Research Foundation (Deutsche Forschungsgemeinschaft DFG; Forschungsgruppe/Research Unit FOR2107). Please see the Supplement for full FOR2107 acknowledgments.

