## Supplementary Material for "Identification of transdiagnostic psychiatric disorder subtypes using unsupervised learning"


**Methods S1.** Individual-level quality control ... 1
**Methods S2.** Genotyping, quality control, and imputation ... 1
**Methods S3.** Calculation of polygenic scores ... 2
**Methods S4.** Clustering analysis ... 2
**Methods S5.** Characterization of clusters ... 3
**Methods S6.** Statistical testing ... 4
**Methods S7.** Assessment of the information gain by PGS ... 4
**Methods S8.** Replication analysis ... 4
**Results S1.** Prediction of diagnoses using cluster labels ... 4
**Table S1.** Characterization of the replication sample and clusters ... 5
**Table S2.** Variables used in the clustering procedure ... 6
**Table S3.** Most important features from the HDDA analysis ... 8
**Table S4.** Characterization of the discovery sample and its clusters regarding variables used in the clustering process ... 9
**Table S5.** Characterization of the discovery sample and its clusters regarding variables not directly used in the clustering procedure ... 11
**Table S6.** MDD subtype analysis ... 12
**Table S7.** *One-vs-all* HDDA classification analysis using only MDD-diagnosed patients ... 13
**Table S8.** Metrics of genetic lasso regularized regression prediction models ... 14
**Table S9.** Significance testing of genetic analyses with the Westfall and Young procedure – *one-vs-all* comparisons ... 15
**Table S10.** Significance testing of genetic analyses with the Westfall and Young procedure – *one-vs-one* comparisons ... 16
**Table S11.** Assessment of the PGS information gain ... 17
**Table S12.** Significance testing with the Westfall and Young procedure – comparison of discovery-replication pairs ... 18
**Table S13.** Characterization of the replication sample and its clusters with variables used in the clustering process ... 19
**Table S14.** Projection of the discovery-stage lasso models to the replication sample ... 22
**Table S15.** Replication – Significance testing with the WY procedure, *one-vs-all* comparisons .. 23
**Table S16.** Replication – Significance testing with the WY procedure, *one-vs-one* comparisons 24
**Table S17.** Bipolar disorder subtypes across clusters. ... 25
**Figure S1.** Population substructure (genetic ancestry components). ... 26
**Figure S2.** Overview of the clustering pipeline ... 28
**Figure S3.** Results from the clustering pipeline ... 29
**Figure S4.** Distribution of PGS for MDD patients ... 30
**Figure S5.** Discovery-stage HDDA models projected to replication sample ... 31
**Full FOR2107 acknowledgements** ... 32
**Supplemental Material References** ... 33

**Methods S1.** Individual-level quality control

*Discovery sample*
Of 1623 individuals eligible for the discovery sample, three were excluded because they withdrew their consent and one for missing diagnostic information from all subsequent analysis.
Next, 47 pairs of relatives were identified by relatedness analysis conducted in PLINK (v1.90b6.2). Of each pair, the individual with the higher genotyping rate was kept in the analyses. Finally, 322 individuals with missing information in any of the 57 variables used for clustering were excluded. The final discovery sample consisted of 1250 individuals, including 590 healthy controls, 477 MDD, 75 BD, 25 SZA, and 53 SCZ patients, and 30 individuals with other diagnoses disorders including anxiety disorders, adjustment disorders, and substance use disorders.

*Replication sample*
Of 855 individuals eligible for the replication dataset, one was excluded because of a withdrawn consent and two for missing diagnostic information. Next, 27 relatives were removed using the same procedure as applied for the discovery sample. Finally, 201 individuals were excluded due to missing information in any of the variables used for clustering. The final replication sample for the analysis consisted of 622 individuals, including 240 healthy controls, 283 MDD, 44 BD, 13 SZA, and 17 SCZ patients, and 25 individuals with other diagnoses including anxiety, adjustment, and substance use disorders.

**Methods S2.** Genotyping, quality control, and imputation
Genotyping was conducted using the Infinium PsychArray BeadChip, as described previously [1]. The quality control (QC) of genetic data was conducted in PLINK v1.90b6.10 [2] and R v3.5.2, as described previously [3]. Pre-imputation QC of genotype data consisted of the following steps:

1. Removal of SNPs with call rates <98% or a minor allele frequency (MAF) <1%
2. Removal of individuals with genotyping rates <98%
3. Removal of sex mismatches
4. Removal of genetic duplicates
5. Removal of cryptic relatives with pi-hat≥12.5
6. Removal of genetic outliers with a distance from the mean of >4 SD in the first eight multidimensional scaling (MDS) ancestry components
7. Removal of individuals with a deviation of the autosomal or X-chromosomal heterozygosity from the mean >4 SD
8. Removal of non-autosomal variants
9. Removal of SNPs with call rates <98% or a MAF <1% or Hardy-Weinberg Equilibrium (HWE) test *p*-values $<1\times10^{-6}$
10. Removal of A/T and G/C SNPs
11. Update of variant IDs and positions to the IDs and positions in the 1000 Genomes Phase 3 reference panel
12. Alignment of alleles to the reference panel
13. Removal of duplicated variants and variants not present in the reference panel

For the calculation of ancestry components (used to determine genetic outliers and as covariates in the analyses), pre-imputation genotype data were used. Additional variant filtering steps were: removal of variants with a MAF <0.05 or HWE *p*-value $<10^{-3}$; removal of variants mapping to the extended MHC region (chromosome 6, 25-35 Mbp) or to a typical inversion site on chromosome 8 (7-13 Mbp); linkage disequilibrium (LD) pruning (command `--indep-pairwise 200 100 0.2`). Next, the pairwise identity-by-state (IBS) matrix of all individuals was calculated using the command `--genome` on the filtered genotype data. Multidimensional scaling (MDS) analysis was performed on the IBS matrix using the eigendecomposition-based algorithm in PLINK v1.90b6.10.

After imputation, variants with a MAF <1%, an HWE test $p<1\times10^{-6}$, and an INFO metric <0.8 were removed. Imputation was conducted using SHAPEIT v2 (r837) [4], IMPUTE2 v2.3.2 [5, 6], and the 1000 Genomes Phase 3 reference panel.
In total, imputed genetic data were available for 1,416 individuals in the discovery and 556 individuals in the replication sample. These individuals were imputed as part of a larger dataset with N=2,248.
Variants before QC: 596,861; variants after QC: 284,691; variants after imputation: 8,565,143.

**Methods S3.** Calculation of polygenic scores

Ten PGSs were calculated for the following disorders and traits, using training summary statistics from published genome-wide association studies (GWASs): ADHD [7] with 19,099 cases and 34,194 controls, ASD [8] with 18,382 cases and 27,969 controls, BD [9] with 20,352 cases and 31,358 controls, psychiatric cross-disorder (CD) [10] with 162,151 cases and 276,846 controls, educational attainment (EA) [11] with 293,723 individuals, extraversion [12] with 63,030 individuals, hedonic well-being (HWB) [13] with 221,575 individuals, MDD [14] with 170,756 cases and 329,443 controls, neuroticism [15] with 329,821 individuals, and SCZ [16] with 40,675 cases and 64,643 controls.

PGSs were calculated using the PRS-CS [17] method that employs Bayesian regression to infer PGS weights while modeling local linkage disequilibrium (LD) patterns using the 1000 Genomes EUR reference panel. All training GWAS variants with an INFO metric <0.6, a MAF <1%, or which were not present in the FOR2107 imputation were removed before estimating PRS-CS weights. The global shrinkage parameter was fixed to $\varphi$=0.01, appropriate for highly polygenic traits. PGSs were calculated in *R* using the PRS-CS weights and imputed dosage data, as described previously [18].

**Methods S4.** Clustering analysis

The clustering analysis was conducted in *R* v3.6.0 using the discovery sample. Clinical variables were scaled and submitted to the HDDC algorithm [19] implemented in the *R* package *HDclassif* [20]. This package implements a subspace clustering algorithm based on the Gaussian mixture model framework, which allowed us to fit 14 different models, corresponding to different regularizations for the cluster solutions, as explained in detail elsewhere [19]. In order to choose the model best fitting for our data, HDDC was run 100 times using cross-validation. In each run, 80% of the discovery dataset was randomly resampled without replacement. The best-fitting model, AkBkQkDk, was chosen using the Integrated Completed Likelihood (ICL) criterion [21].

After the choice of the optimal model (AkBkQkDk), the optimal number of clusters was determined using a leave-one-out jackknife resampling procedure, testing the potential clustering solutions from K=2 to K=15 clusters. In this procedure, the model was fitted 1250 times to the discovery sample containing N=1250 individuals, each time leaving out one individual from the sample. Hence, 1250 clustering solutions were generated for each analyzed K value. The cluster solution where the parameter K reached the best median ICL value in the resampling procedure was selected as the final model (K=5). The final assignment of individuals to clusters for the optimal model AkBkQkDk and parameter K=5 was performed based on the 1250 leave-one-out cluster solutions by using the majority voting scheme, as implemented in the *R* package *dicer* [22]. The stability of the clustering solution was assessed with an additional 100 runs of the model-fitting procedure, each time using 95% of the resampled discovery sample. The thus obtained solution was compared to the previously determined optimal solution using the Rand and Jaccard similarity indices. The illustration of the complete clustering workflow is shown in Figure S2*,* while the results of each step provided in Figure S3.

**Methods S5.** Characterization of clusters

*High-dimensional discriminant analysis*

Supervised high-dimensional discriminant analysis (HDDA) [20, 23] is a subspace classification algorithm that incorporates the same Gaussian mixture model fitting for the high-dimensional data as the corresponding unsupervised HDDC does. In this analysis, the same 57 variables and the best-fitting model identified by HDDC (AkBkQkDk, K=5) were used to predict the cluster labels. The prediction model was run 100 times on the discovery sample, each time on a separate random 70% *vs.* 30% split of training and test data, respectively. For each run, the area under the receiver-operator curve (AUC) was calculated using the *R* package *pROC* [24]. The average metrics across the 100 runs were used as the final measure of classification success.

The importance of variables was assessed to identify the relevant variables for distinguishing each cluster from the others. This importance was measured by calculating the decrease in the model's AUC metric after the values of the respective variable were randomly permuted. The larger the drop of AUC after permutation, the more relevant the variable. The variables were ranked using the average AUC drop after 100 runs.

*Lasso regularized regression*

Lasso (least absolute shrinkage and selection operator) regularized regression [25] analyses were conducted using the *R* package *glmnet* [26], with 24 predictors (10 PGSs, 4 self-reported family history variables, 8 ancestry components, age, and gender). Lasso penalizes regression coefficients and may shrink them to zero; the degree of penalization depends on the parameter $\lambda$. To tune $\lambda$, the discovery sample was split 1000 times into 70% training and 30% test data, using stratification based on cluster labels. On each of the 1000 training sets, $\lambda$ was tuned via three-fold cross-validation. In each run, the $\lambda$ minimizing the cross-validation error, *i.e.*, maximizing the AUC ($\lambda_{min}$), was used to obtain Lasso coefficients on the test set. AUC, sensitivity, and specificity were calculated in each run; the average metrics of all runs were reported. The cutoff for the sensitivity and specificity was determined using the *MaxSpSe* method of the *OptimalCutpoints* package [27]. Lastly, to get the final model, *i.e.*, variable sets and their corresponding coefficients for the cluster prediction, lasso was fit to the full dataset with $\lambda$ equal to the median value of all $\lambda_{min}$ chosen during the tuning procedure.

**Methods S6.** Statistical testing
The Westfall and Young method for significance testing controls the family-wise error rate while accounting for the possible dependence structure of the analyzed variables. The adjusted *p*-values were directly estimated via 20,000 permutations, using the *mt.minP* function from the Bioconductor *multtest* package [28] with Welch's t statistic. After adjusting the *p*-values using the Westfall and Young procedure, they were, subsequently, also corrected for the number of comparisons made using Bonferroni's method. This number of comparisons corresponded to the number of clusters in the *one-vs-all* analyses and to *(K*(K-1))/2* for *one-vs-one* (pairwise) analyses.

**Methods S7.** Assessment of the information gain by PGS
For the assessment of the information gain of PGSs in the prediction of clusters, we analyzed four multinomial regression models using the *R* package VGAM [29] with cluster labels as the dependent variable and the following predictors: a) PGS and ancestry components (ACs), b) family history only, c) family history and ACs, d) the full model with PGS, family history, and ACs. Age and gender were used as covariates in all four models. The model performance was assessed using Nagelkerke's pseudo-$R^2$, the Akaike information criterion (AIC), and likelihood-ratio tests. For the calculation of the pseudo-$R^2$, a null model containing only the covariates age and gender was used.

**Methods S8.** Replication analysis
To identify the discovery-stage clusters in the replication data, each of the binary discovery-stage *one-vs-all* HDDA classification models was used for *one-vs-all* predictions in the replication dataset, thereby conducting $K^2$ comparisons. The discovery-stage model producing the best prediction above a threshold of 70% was chosen to assign the cluster identity in the replication sample. Further statistical analysis, *i.e.*, significance testing with the Westfall and Young procedure (described in the Methods S6) was carried out to confirm the matching and homogeneity of linked discovery and replication clusters. As for the discovery sample, *p*-values adjusted by the Westfall and Young procedure were further corrected using Bonferroni's method for the five comparisons made. Genetic prediction analyses were conducted as described above, using the lasso model with the λ parameter optimized in the discovery sample.

**Results S1.** Prediction of diagnoses using cluster labels
Our cluster solution crossed diagnostic boundaries mostly for MDD and BD, whereas cluster 4 was enriched for SCZ and SZA patients (Table 1, Figure 1A). Using multinomial regression models, we predicted DSM-IV diagnoses with our cluster labels. When adding cluster labels to a null model containing only age and gender, Nagelkerke's pseudo-$R^2$ increased by 42%.

**Table S1.** Characterization of the replication sample and clusters

We compared age, sex, and diagnosis of the discovery and replication datasets. Age and diagnosis differed significantly (*p*=0.01 and *p*=0.002, respectively), while sex did not (*p*=0.16). Proportions of single diagnostic categories were significant for healthy controls (p=0.005) and MDD (p-value 0.003). Other categories did not show significant differences (BD, *p*=0.4; SCZ, *p*=0.1; SZA, *p*=1; Other diagnosis, *p*=0.07). Age at Onset was defined according to OPCRIT item 4 and was not available for HC.
**Abbreviations and clarifications:** BMI = Body mass index; Family history Any = Family history of any psychiatric disorder; AAO = Age at Onset (*not available for healthy controls); HC = Healthy controls; BD = Bipolar Disorder; MDD = Major Depressive Disorder; SCZ = Schizophrenia; SZA = Schizoaffective Disorder; Other = other psychiatric diagnoses including anxiety, adjustment, and substance use disorders; SANS/SAPS = Scale for the Assessment of Negative/Positive Symptoms; SF36; 36-Item Short Form Survey.
**Variable ranges and interpretations**: Quality of life [0-100, 0=disability]; HAMD 21 item [0-66, 0=not present]; HAMA [0-56, 0=not present]; BDI [0-63, 0=not present]; SAPS/SANS [0-86/0-80, 0=not present]; CTQ [5-25, 5=less severe experiences].

| Variable | Full | Cluster 0 | Cluster 1 | Cluster 2 | Cluster 3 | Cluster 4 |
|---|---|---|---|---|---|---|
| N | 622 | 128 | 178 | 59 | 136 | 121 |
| Demographics | | | | | | |
| Age, mean (SD) | 36.3 (12.6) | 34 (11.5) | 38.8 (13.3) | 31.6 (10.7) | 36.1 (13.0) | 37.8 (12.2) |
| Gender, N(%) male | 219 (35%) | 41 (32%) | 65 (37%) | 18 (31%) | 51 (38%) | 44 (36%) |
| Years of education, mean (SD) | 13.8 (2.8) | 15 (2.6) | 14.1 (2.7) | 13.4 (2.2) | 13.5 (2.5) | 12.6 (2.9) |
| Living with partner, N (%) | 199 (33%) | 47 (37%) | 67 (38%) | 13 (23%) | 39 (29%) | 33 (29%) |
| BMI, mean (SD) | 25.6 (5.4) | 23.3 (3.4) | 25.8 (5.5) | 24.9 (5.3) | 26 (4.8) | 27.5 (6.9) |
| Family history Any, N (%) | 306 (50%) | 54 (42%) | 83 (48%) | 33 (58%) | 66 (50%) | 70 (61%) |
| Diagnosis | | | | | | |
| AAO*, mean (SD) | 24.1 (11.7) | 24.3 (10.4) | 23.2 (9.9) | 20.1 (9.8) | 26.5 (12.5) | 23.6 (12.3) |
| HC, N (%) | 240 (39%) | 104 (81%) | 96 (54%) | 11 (19%) | 17 (13%) | 12 (10%) |
| BD, N (%) | 44 (7%) | 1 (1%) | 9 (5%) | 7 (12%) | 17 (13%) | 10 (8%) |
| MDD, N (%) | 283 (45%) | 17 (13%) | 60 (34%) | 38 (64%) | 88 (65%) | 80 (66%) |
| SCZ, N (%) | 17 (3%) | 0 (0%) | 3 (2%) | 1 (2%) | 5 (4%) | 8 (7%) |
| SZA, N (%) | 13 (2%) | 0 (0%) | 2 (1%) | 1 (2%) | 1 (1%) | 9 (7%) |
| Other, N (%) | 25 (4%) | 6 (5%) | 8 (4%) | 1 (2%) | 8 (6%) | 2 (2%) |
| Quality of life (SF36), mean (SD) | | | | | | |
| General health | 68.1 (23) | 88.6 (12.8) | 75.9 (16.3) | 64.5 (16.6) | 56.3 (20.8) | 49.8 (22.4) |
| Mental health | 62.8 (22.1) | 83.7 (9.4) | 73.4 (13.8) | 55.1 (16.8) | 48.2 (17.2) | 45.5 (22.0) |
| Depression and anxiety, mean (SD) | | | | | | |
| HAMA sum | 8.4 (8.2) | 2.5 (3) | 4.8 (4.5) | 9.7 (6.8) | 11.6 (7.5) | 15.7 (10.2) |
| HAMD sum | 6.1 (6.8) | 1.3 (2.1) | 2.9 (3.2) | 7.4 (6.4) | 8.8 (6.5) | 12.2 (7.9) |
| BDI sum | 11.3 (10.3) | 2.9 (2.8) | 6.6 (5.7) | 14.6 (8.4) | 15.8 (9.1) | 20.4 (12.1) |
| Positive and negative Symptoms, mean (SD) | | | | | | |
| SANS | 4.3 (7.3) | 0.3 (0.9) | 1.7 (3.1) | 4 (5.6) | 6.4 (7.3) | 10.4 (10.7) |
| SAPS | 0.7 (2.7) | 0.05 (0.2) | 0.2 (0.6) | 0.5 (1.4) | 0.4 (1.2) | 2.8 (5.3) |
| Maltreatment in Childhood and Youth (CTQ), mean (SD) | | | | | | |
| Emotional abuse | 9.1 (4.5) | 6.1 (1.5) | 8.2 (2.8) | 13.4 (4.8) | 7.8 (3) | 13.2 (5.5) |
| Emotional neglect | 11.1 (5.2) | 6.9 (2.1) | 10.8 (3.7) | 16.6 (4.7) | 9.4 (3.4) | 15 (6.1) |
| Physical abuse | 6.1 (2.4) | 5.2 (0.6) | 5.7 (1.3) | 6 (1.7) | 5.5 (1) | 8.4 (4.1) |
| Physical neglect | 6.8 (2.5) | 5.4 (1.0) | 6.5 (1.4) | 8.5 (2.3) | 6.1 (1.7) | 8.8 (3.6) |
| Sexual abuse | 5.7 (2.5) | 5 (0.3) | 5.3 (1) | 5.3 (1.0) | 5.3 (1.2) | 7.8 (4.8) |

**Table S2.** Variables used in the clustering procedure

The values represent mean (SD) of the 57 variables used in the clustering procedure for the discovery (N=1250) and replication (N=622) sample.

| Category | Variable | Discovery | Replication |
|---|---|---|---|
| Attachment style / Relationship Scales Questionnaire | RSQ Anxiety of separation | 2.7 (0.7) | 2.7 (0.7) |
| | RSQ Avoidance of closeness | 2.5 (0.9) | 2.5 (0.9) |
| | RSQ Desire for independence | 3.9 (0.8) | 3.97 (0.7) |
| | RSQ Lack of trust | 2.3 (0.9) | 2.4 (0.9) |
| Depression and anxiety level | BDI-II Sum | 10.7 (10.8) | 11.3 (10.3) |
| | HAMA Sum | 7.3 (7.9) | 8.4 (8.2) |
| | HAMD Sum21 | 5.4 (6.6) | 6.1 (6.8) |
| | STAIS | 42.2 (13.2) | 43.0 (12.8) |
| | STAIT | 43.2 (14.2) | 44.2 (13.8) |
| Anhedonia | SHAPS | 1.99 (2.9) | 1.97 (2.7) |
| Life events | LEQ Negative Events score | 10.0 (13.8) | 10.4 (12.3) |
| | LEQ Positive Events score | 9.8 (9.3) | 9.4 (9.3) |
| Life stress | PSS Sum | 22.8 (10.8) | 23.8 (10.5) |
| Maltreatment in childhood and youth | ACE Sum | 1.6 (1.9) | 1.6 (1.8) |
| | CTQ Emotional abuse | 9.1 (4.7) | 9.1 (4.5) |
| | CTQ Emotional neglect | 10.7 (5.2) | 11.1 (5.2) |
| | CTQ Physical abuse | 6.2 (2.6) | 6.1 (2.4) |
| | CTQ Physical neglect | 7.2 (2.7) | 6.8 (2.5) |
| | CTQ Sexual abuse | 5.8 (2.5) | 5.7 (2.5) |
| Mania Symptoms | YMRS | 1.2 (2.5) | 1.4 (2.7) |
| Negative symptoms | SANS sum score | 5.7 (9.9) | 4.3 (7.3) |
| Positive symptoms | SAPS sum score | 1.4 (5.2) | 0.7 (2.7) |
| Personality | NEO-FFI Agreeableness | 33.1 (6.0) | 33.2 (6.1) |
| | NEO-FFI Conscientiousness | 32 (7.5) | 32.2 (7.4) |
| | NEO-FFI Extraversion | 26.4 (8.2) | 25.8 (8.2) |
| | NEO-FFI Neuroticism | 22.1 (10.5) | 23.1 (10.2) |
| | NEO-FFI Openness to experience | 30.3 (6.96) | 30.4 (6.8) |
| | SPQB (Schizotypy) | 5.95 (4.7) | 5.97 (4.7) |
| Protective factors | Maternal bonding | 24.7 (8.6) | 24.4 (8.5) |
| | Paternal bonding | 22.3 (8.7) | 21.2 (8.9) |
| | RS25 Sum Score (Resilience) | 125.4 (27.2) | 125.1 (26.9) |
| | Social support | 4.1 (0.8) | 4.1 (0.7) |
| Short Form Health Survey / Quality of life | SF36 Bodily pain | 76.3 (26.2) | 74.7 (26.1) |
| | SF36 Energy/fatigue | 50.2 (23) | 47.5 (22.1) |
| | SF36 General health | 65.96 (23.4) | 68.1 (23) |

| | | | |
|---|---|---|---|
| | SF36 Mental health | 64.3 (22.6) | 62.8 (22.1) |
| | SF36 Physical functioning | 89.5 (17.1) | 89.8 (15.5) |
| | SF36 Role emotional | 65.5 (42.4) | 60.6 (43.2) |
| | SF36 Role physical | 76.6 (36.7) | 71.2 (36.1) |
| | SF36 Social functioning | 73.3 (29.95) | 70.4 (30.1) |
| Symptom Checklist | SCL90R Additional Items | 4.1 (4.0) | 4.4 (3.8) |
| | SCL90R Anxiety | 5.4 (6.7) | 5.4 (6.2) |
| | SCL90R Depression | 11.4 (11.9) | 12.2 (11.6) |
| | SCL90R Global severity index | 0.6 (0.6) | 0.6 (0.5) |
| | SCL90R Hostility | 2.9 (3.7) | 3.2 (3.9) |
| | SCL90R Interpersonal sensitivity | 6.8 (7.3) | 6.96 (7.0) |
| | SCL90R Obsessive–compulsive behavior | 8.4 (8.1) | 8.7 (7.7) |
| | SCL90R Paranoid ideation | 3.5 (4.4) | 3.3 (4.0) |
| | SCL90R Phobic anxiety | 2.3 (4.1) | 2.3 (3.9) |
| | SCL90R Positive symptom distress Index | 1.5 (0.5) | 1.5 (0.5) |
| | SCL90R Positive symptom total | 29.8 (21.0) | 31.2 (19.5) |
| | SCL90R Psychoticism | 3.9 (5.2) | 3.7 (4.9) |
| | SCL90R Somatization | 6.96 (7.1) | 7.1 (6.5) |
| Verbal IQ | Verbal IQ | 113.99 (13.7) | 113.8 (13.4) |
| Verbal Learning and Memory Test | VLMT Sum | 56.8 (10.2) | 56.1 (9.6) |
| Visuospatial Working Memory Test | Corsi block-tapping test | 17.3 (3.4) | 17.2 (3.1) |
| Visuospatial Working Memory Test | Letter Number Span test | 16.1 (3.2) | 16.1 (3.4) |

**Table S3.** Most important features from the HDDA analysis

The most important variables for each cluster from the '*one-vs.-all'* HDDA classification analysis. The importance was calculated based on the average AUC drop, as explained in the Methods S5.

| Cluster 0 | Cluster 1 | Cluster 2 | Cluster 3 | Cluster 4 |
|---|---|---|---|---|
| CTQ Sexual abuse | SF36 Bodily pain | Maternal bonding, care | SF36 Role physical | SAPS (Positive symptoms) |
| LEQ Negative Events score | SF36 Role physical | CTQ Emotional neglect | CTQ Emotional neglect | YMRS |
| SAPS (Positive Symptoms) | NEO-FFI Agreeableness | Paternal bonding, care | Maternal bonding, care | CTQ Physical abuse |
| CTQ Physical abuse | RSQ Lack of trust | SF36 Physical functioning | CTQ Emotional abuse | CTQ Sexual abuse |
| SF36 Physical functioning | Maternal bonding, care | SF36 Role physical | CTQ Physical neglect | LEQ Negative Events score |
| YMRS | STAIS | ACE Sum | ACE Sum | SANS (Negative symptoms) |
| SANS (Negative symptoms) | SHAPS | RSQ Avoidance of closeness | SF36 Physical functioning | CTQ Physical neglect |
| SF36 Role physical | SF36 Physical functioning | CTQ Emotional abuse | CTQ Physical abuse | LEQ Positive Events score |
| SCL90R Phobic anxiety | NEO-FFI Neuroticism | CTQ Physical abuse | SF36 Energy | VLMT Sum |
| CTQ Physical neglect | SAPS (Positive symptoms) | SAPS (Positive Symptoms) | Paternal bonding, care | CTQ Emotional abuse |

**Table S4.** Characterization of the discovery sample and its clusters regarding variables used in the clustering process

| Category | Variable, mean (SD) | Full | Cluster 0 | Cluster 1 | Cluster 2 | Cluster 3 | Cluster 4 |
|---|---|---|---|---|---|---|---|
| | N | 1250 | 535 | 38 | 266 | 215 | 196 |
| Attachment style / Relationship Scales Questionnaire | RSQ Anxiety of separation | 2.7 (0.7) | 2.5 (0.5) | 2.8 (0.8) | 2.9 (0.8) | 2.9 (0.8) | 2.8 (0.8) |
| | RSQ Avoidance of closeness | 2.5 (0.9) | 1.98 (0.6) | 2.7 (0.9) | 2.9 (0.8) | 2.5 (0.8) | 3.1 (0.9) |
| | RSQ Desire for independence | 3.9 (0.8) | 3.8 (0.8) | 4.0 (0.7) | 4.1 (0.7) | 3.8 (0.9) | 4.0 (0.8) |
| | RSQ Lack of trust | 2.3 (0.9) | 1.8 (0.6) | 2.8 (0.9) | 2.7 (0.8) | 2.5 (0.9) | 3.0 (0.9) |
| Depression and anxiety level | BDI-II Sum | 10.7 (10.8) | 3.2 (3.3) | 15.1 (12.8) | 12.7 (9.6) | 17.6 (10.1) | 20.3 (11.7) |
| | HAMA Sum | 7.3 (7.9) | 2.2 (2.5) | 9.7 (8.6) | 7.6 (6.4) | 13.1 (8.6) | 14.0 (8.8) |
| | HAMD Sum21 | 5.4 (6.6) | 1.1 (1.6) | 6.3 (6.7) | 5.9 (5.7) | 9.9 (7.0) | 10.95 (7.5) |
| | STAIS | 42.2 (13.2) | 33 (6.4) | 48.9 (12.2) | 44.9 (11.7) | 51.7 (13.1) | 51.5 (13.1) |
| | STAIT | 43.2 (14.2) | 32.6 (6.9) | 49.3 (14.1) | 47.6 (12.8) | 53.1 (12.5) | 54.2 (12.5) |
| Anhedonia | SHAPS | 1.99 (2.9) | 0.6 (1.2) | 2.6 (4.0) | 2.3 (2.7) | 3.5 (3.5) | 3.6 (3.7) |
| Life events | LEQ Negative Events score | 10 (13.8) | 3.3 (4.3) | 12.4 (11.8) | 9.99 (8.9) | 14.2 (11.1) | 23.4 (24.1) |
| | LEQ Positive Events score | 9.8 (9.3) | 9.4 (7.1) | 8.1 (7.7) | 9.5 (8.4) | 8.7 (9.5) | 12.5 (14) |
| Life stress | PSS Sum | 22.8 (10.8) | 15 (6.1) | 26.7 (10.8) | 25.6 (8.9) | 31.1 (9.7) | 30.6 (9.6) |
| Maltreatment in childhood and youth | ACE Sum | 1.6 (1.9) | 0.5 (0.8) | 1.7 (1.6) | 2.4 (1.7) | 1.3 (1.3) | 3.96 (2.4) |
| | CTQ Emotional abuse | 9.1 (4.7) | 6.3 (1.7) | 9.9 (4.9) | 11.4 (4.2) | 7.99 (3.0) | 14.6 (5.9) |
| | CTQ Emotional neglect | 10.7 (5.2) | 7.5 (2.6) | 11.8 (5.4) | 14.1 (4.1) | 9.2 (3.5) | 16.5 (5.4) |
| | CTQ Physical abuse | 6.2 (2.6) | 5.3 (0.7) | 6.4 (2.2) | 6.4 (2) | 5.5 (1.1) | 9.5 (4.6) |
| | CTQ Physical neglect | 7.2 (2.7) | 5.8 (1.4) | 7.2 (2.2) | 8 (2.2) | 6.4 (1.6) | 10.4 (3.7) |
| | CTQ Sexual abuse | 5.8 (2.5) | 5.1 (0.4) | 5.8 (2.0) | 5.8 (1.9) | 5.6 (1.7) | 7.98 (4.9) |
| Mania Symptoms | YMRS | 1.2 (2.5) | 0.5 (1.1) | 0.8 (1.4) | 1.1 (1.8) | 1.3 (2) | 2.9 (4.9) |
| Negative symptoms | SANS sum score | 5.7 (9.9) | 0.6 (1.7) | 5.2 (8.6) | 6.9 (9.6) | 7.96 (9.1) | 15.6 (14.4) |
| Positive symptoms | SAPS sum score | 1.4 (5.2) | 0.1 (0.6) | 0.1 (0.4) | 0.6 (1.6) | 0.7 (1.8) | 6.7 (11.5) |
| Personality | NEO-FFI Agreeableness | 33.1 (6.0) | 35.6 (5.1) | 30.5 (4.8) | 31.1 (5.5) | 32.9 (6.3) | 29.5 (5.9) |
| | NEO-FFI Conscientiousness | 32 (7.5) | 34.9 (6.1) | 30.1 (6.9) | 29.4 (7.7) | 30.7 (7.5) | 29.4 (7.8) |
| | NEO-FFI Extraversion | 26.4 (8.2) | 30.8 (6.1) | 23.7 (9.2) | 23.5 (8) | 24.2 (7.3) | 21.1 (7.9) |
| | NEO-FFI Neuroticism | 22.1 (10.5) | 14.7 (6.6) | 26.9 (9.5) | 26 (9.4) | 28.2 (9.1) | 29.4 (9.1) |
| | NEO-FFI Openness to experience | 30.3 (7.0) | 31.2 (6.6) | 29.1 (5.2) | 31.1 (6.9) | 28.8 (7.4) | 28.6 (7.4) |
| | SPQB (Schizotypy) | 5.95 (4.7) | 2.9 (2.6) | 7.0 (4.7) | 7.95 (4.2) | 6.6 (4.3) | 10.6 (4.8) |

| Category | Variable | Full | Cluster 0 | Cluster 1 | Cluster 2 | Cluster 3 | Cluster 4 |
|---|---|---|---|---|---|---|---|
| Protective factors | Maternal bonding, care | 24.7 (8.6) | 29.8 (4.8) | 21.6 (9.9) | 19.4 (7.4) | 27 (6.2) | 15.7 (8.7) |
| | Paternal bonding, care | 22.3 (8.7) | 26.95 (6.3) | 22.5 (9.2) | 16.9 (7.4) | 23.2 (7.6) | 15.7 (8.7) |
| | RS25 Sum Score (Resilience) | 125.4 (27.2) | 140.9 (16.8) | 119.9 (25.1) | 117.9 (26) | 112.6 (27.3) | 108.6 (30.1) |
| | Social support | 4.1 (0.8) | 4.6 (0.4) | 3.8 (1.0) | 3.8 (0.8) | 4.0 (0.8) | 3.5 (0.96) |
| Short Form Health Survey / Quality of life | SF36 Bodily pain | 76.3 (26.2) | 88.9 (15.6) | 53.9 (23.9) | 78.9 (22.8) | 61.1 (29.4) | 59.7 (30) |
| | SF36 Energy/fatigue | 50.2 (23.0) | 66.5 (13.0) | 40 (25.4) | 45.9 (19.3) | 31.2 (18.5) | 34.1 (22.2) |
| | SF36 General health | 66 (23.4) | 81.2 (14.0) | 57.9 (26.6) | 64.7 (19.8) | 49.4 (20.7) | 45.8 (21.3) |
| | SF36 Mental health | 64.3 (22.6) | 81.4 (9.8) | 55.6 (23) | 59.4 (18.9) | 46.4 (19.3) | 45.6 (21.3) |
| | SF36 Physical functioning | 89.5 (17.1) | 97.6 (4.9) | 81.97 (19.95) | 93.3 (9.1) | 78.2 (23.2) | 76.3 (22.5) |
| | SF36 Role emotional | 65.5 (42.4) | 94.6 (16.4) | 56.1 (43.2) | 58.4 (41.3) | 31.5 (39.4) | 34.9 (42.3) |
| | SF36 Role physical | 76.6 (36.7) | 96.8 (11.5) | 52 (39.2) | 85.1 (27.9) | 44.8 (40.9) | 49.5 (43.5) |
| | SF36 Social functioning | 73.3 (30) | 95.2 (10) | 66.4 (31) | 67.6 (26.0) | 49.4 (28.2) | 48.5 (29.5) |
| Symptom checklist | SCL90R Additional Items | 4.1 (4.0) | 1.6 (1.7) | 5.1 (3.8) | 4.2 (3.2) | 6.9 (4.4) | 7.7 (4.3) |
| | SCL90R Anxiety | 5.4 (6.7) | 1.3 (1.7) | 7.6 (6.9) | 5.5 (4.96) | 9.6 (7.3) | 11.3 (8.6) |
| | SCL90R Depression | 11.4 (11.9) | 2.7 (2.7) | 15.7 (13.8) | 13.4 (10.0) | 20.4 (11.7) | 21.7 (12.4) |
| | SCL90R Global severity index | 0.6 (0.6) | 0.2 (0.1) | 0.8 (0.6) | 0.6 (0.4) | 0.98 (0.6) | 1.2 (0.6) |
| | SCL90R Hostility | 2.9 (3.7) | 0.8 (1.1) | 4.1 (4.2) | 3.0 (2.7) | 4.9 (4.2) | 5.8 (5.1) |
| | SCL90R Interpersonal sensitivity | 6.8 (7.3) | 1.9 (2.1) | 8.6 (8.1) | 8.8 (6.4) | 10.3 (7.1) | 13.8 (8.4) |
| Symptom checklist | SCL90R Obsessive–compulsive behavior | 8.4 (8.1) | 2.7 (2.4) | 10.1 (8.0) | 9.5 (6.6) | 14.6 (8.3) | 15.3 (8.4) |
| | SCL90R Paranoid ideation | 3.5 (4.4) | 0.8 (1.4) | 4.2 (4.7) | 4.1 (3.5) | 4.9 (4.6) | 8.1 (5.3) |
| | SCL90R Phobic anxiety | 2.3 (4.1) | 0.3 (0.8) | 2.4 (3.3) | 2.1 (2.7) | 3.9 (4.6) | 6.4 (6.3) |
| | SCL90R Positive symptom distress Index | 1.5 (0.5) | 1.1 (0.2) | 1.7 (0.5) | 1.5 (0.4) | 1.9 (0.5) | 1.98 (0.6) |
| | SCL90R Positive symptom total | 29.8 (21.0) | 12.8 (8.7) | 37.0 (20.7) | 35.9 (16.7) | 44.4 (17.4) | 50.4 (17.7) |
| | SCL90R Psychoticism | 3.9 (5.2) | 0.7 (1.1) | 4.9 (5.2) | 4.3 (3.9) | 6.2 (5.1) | 9.4 (7.0) |
| | SCL90R Somatization | 6.96 (7.1) | 2.8 (2.4) | 10.3 (7.4) | 6.3 (4.6) | 12.1 (7.9) | 12.8 (9.3) |
| Verbal IQ | Verbal IQ | 114 (13.7) | 114.8 (13.6) | 116.1 (12.2) | 115.7 (13.9) | 112.4 (12.8) | 110.8 (14.5) |
| Verbal Learning and memory test | VLMT Sum | 56.8 (10.2) | 60.0 (8.6) | 57.3 (9.5) | 56.95 (8.7) | 55.99 (9.9) | 48.7 (11.7) |
| Visuospatial Working Memory Test | Corsi block-tapping test | 17.3 (3.4) | 18.4 (3.2) | 17.2 (3.4) | 17.1 (3.0) | 16.7 (3.2) | 15.3 (3.6) |
| Visuospatial Working Memory Test | Letter Number Span test | 16.1 (3.2) | 16.8 (3.1) | 16.4 (2.9) | 16.5 (2.8) | 15.9 (3) | 14 (3.4) |

**Table S5.** Characterization of the discovery sample and its clusters regarding variables not directly used in the clustering procedure

SANS and SAPS variables were used in the clustering process as sum scores. Here, we present the subscales. Medication, smoking and sociodemographic were not used in the clustering.

| Variable | Full | Cluster 0 | Cluster 1 | Cluster 2 | Cluster 3 | Cluster 4 |
|---|---|---|---|---|---|---|
| N | 1250 | 535 | 38 | 266 | 215 | 196 |
| Positive Symptoms (SAPS) | | | | | | |
| Hallucinations, mean (SD) | 0.3 (1.8) | 0.03 (0.21) | 0 (0) | 0.03 (0.23) | 0.07 (0.55) | 1.49 (4.24) |
| Bizarre Behavior, mean (SD) | 0.09 (0.7) | 0.01 (0.13) | 0 (0) | 0.06 (0.31) | 0.08 (0.53) | 0.41 (1.53) |
| Positive Formal Thought Disorder, mean (SD) | 0.5 (1.9) | 0.08 (0.43) | 0.05 (0.32) | 0.29 (1.1) | 0.5 (1.4) | 2.33 (4.13) |
| Delusions, mean (SD) | 0.5 (2.5) | 0.02 (0.15) | 0.05 (0.23) | 0.19 (0.87) | 0.09 (0.49) | 2.48 (5.75) |
| Negative Symptoms (SANS) | | | | | | |
| Anhedonia, mean (SD) | 1.9 (3.5) | 0.1 (0.7) | 2.1 (3.5) | 2.3 (3.4) | 3.1 (4.1) | 4.5 (4.9) |
| Affective blunting, mean (SD) | 1.5 (3.6) | 0.3 (0.9) | 1.4 (2.9) | 1.83 (3.9) | 1.72 (3.26) | 4.2 (5.9) |
| Avolition / Apathy, mean (SD) | 1.3 (2.5) | 0.09 (0.47) | 0.8 (1.8) | 1.4 (2.4) | 1.8 (2.4) | 3.8 (3.7) |
| Alogia, mean (SD) | 0.5 (1.6) | 0.07 (0.4) | 0.2 (0.9) | 0.7 (1.8) | 0.6 (1.5) | 1.5 (2.8) |
| Medication | | | | | | |
| Antidepressant, yes, N (%) | 350 (28%) | 23 (4%) | 12 (32%) | 94 (35%) | 122 (57%) | 99 (51%) |
| Antipsychotic, yes, N (%) | 187 (15%) | 12 (2%) | 4 (11%) | 41 (15%) | 46 (21%) | 84 (43%) |
| Mood stabilizer, yes, N (%) | 65 (5%) | 9 (2%) | 1 (3%) | 7 (3%) | 22 (10%) | 26 (13%) |
| Antidepressant + Antipsychotic, yes, N (%) | 111 (9%) | 3 (1%) | 2 (5%) | 29 (11%) | 35 (16%) | 42 (21%) |
| Antidepressant + Mood stabilizer, yes, N (%) | 40 (3%) | 4 (1%) | 1 (3%) | 3 (1%) | 16 (7%) | 16 (8%) |
| Antipsychotic + Mood stabilizer, yes, N (%) | 31 (2%) | 3 (1%) | 1 (3%) | 4 (2%) | 9 (4%) | 14 (7%) |
| Smoking | | | | | | |
| No or minimal addiction, N (%) | 1071 (86%) | 498 (93%) | 34 (89%) | 228 (86%) | 175 (81%) | 136 (69%) |
| Average addiction, N (%) | 68 (5%) | 21 (4%) | 1 (3%) | 19 (7%) | 8 (4%) | 19 (10%) |
| Strong addiction, N (%) | 70 (6%) | 10 (2%) | 1 (3%) | 16 (6%) | 22 (10%) | 21 (11%) |
| Very strong addiction, N (%) | 41 (3%) | 6 (1%) | 2 (5%) | 3 (1%) | 10 (5%) | 20 (10%) |
| Sociodemographic - Type of living | | | | | | |
| Alone, N (%) | 342 (27%) | 105 (20%) | 16 (42%) | 90 (34%) | 64 (30%) | 67 (34%) |
| Marriage/life partner, N (%) | 277 (22%) | 108 (20%) | 7 (18%) | 51 (19%) | 62 (29%) | 49 (25%) |
| Parents/relatives, N (%) | 128 (10%) | 45 (8%) | 3 (8%) | 29 (11%) | 27 (13%) | 24 (12%) |
| Non-martial partner, N (%) | 167 (13%) | 89 (17%) | 4 (11%) | 29 (11%) | 21 (10%) | 24 (12%) |
| Therapeutic facilities, N (%) | 12 (1%) | 0 (0%) | 0 (0%) | 0 (0%) | 1 (0%) | 11 (6%) |
| Shared flat, N (%) | 303 (24%) | 185 (35%) | 7 (18%) | 61 (23%) | 31 (14%) | 19 (10%) |
| Other, N (%) | 15 (1%) | 2 (0%) | 1 (3%) | 4 (2%) | 6 (3%) | 2 (1%) |
| Sociodemographic - Income | | | | | | |
| Own work, N (%) | 541 (43%) | 259 (48%) | 16 (42%) | 117 (44%) | 87 (40%) | 62 (32%) |
| Parent/Partner/other, N (%) | 334 (27%) | 201 (38%) | 10 (26%) | 53 (20%) | 48 (22%) | 22 (11%) |
| Wage replacement / Sickness pay / Unemployment pay, N (%) | 276 (22%) | 35 (7%) | 11 (29%) | 70 (26%) | 60 (28%) | 100 (51%) |
| Other, N (%) | 86 (7%) | 35 (7%) | 1 (3%) | 24 (9%) | 17 (8%) | 9 (5%) |
| Sociodemographic - Social contacts | | | | | | |
| 1 time per week, N (%) | 274 (22%) | 97 (18%) | 10 (26%) | 64 (24%) | 60 (28%) | 43 (22%) |
| Several times per week, N (%) | 722 (58%) | 402 (75%) | 14 (37%) | 139 (52%) | 90 (42%) | 77 (39%) |
| 1 time every 14 days, N (%) | 112 (9%) | 20 (4%) | 4 (11%) | 31 (12%) | 31 (14%) | 26 (13%) |
| 1 time in month, including distant acquaintances, N (%) | 95 (8%) | 10 (2%) | 7 (18%) | 27 (10%) | 22 (10%) | 29 (15%) |
| No social contact apart from meeting at work, N (%) | 35 (3%) | 4 (1%) | 3 (8%) | 4 (2%) | 11 (5%) | 13 (7%) |
| Meeting friends under no circumstances, N (%) | 7 (1%) | 0 (0%) | 0 (0%) | 0 (0%) | 0 (0%) | 7 (4%) |

**Table S6.** MDD subtype analysis

This table shows phenotypic characteristics of MDD patients, constituting a secondary, descriptive analysis to assess the heterogeneity within MDD patients. The significance of genetic variables was tested using the Westfall and Young procedure (Methods S6). Here, only the significant variables are shown. The ADHD PGS (*) was not significant in the significance analysis of the full sample including all diagnoses. The distributions of the two significant PGS per cluster are shown in Figure S4 A-B. The age at onset was defined according to OPCRIT item 4.
Abbreviations: EA, educational attainment; ADHD, Attention deficit/hyperactivity disorder, W-Y, Westfall and Young.

| Variable | Discovery MDD | Cluster 0 | Cluster 1 | Cluster 2 | Cluster 3 | Cluster 4 |
|---|---|---|---|---|---|---|
| N (MDD diagnosis) | 477 | 56 | 17 | 152 | 147 | 105 |
| Age at Onset, mean (SD) | 25.8 (12.5) | 25.3 (11.1) | 29.1 (13.2) | 23.7 (12.0) | 28.1 (13) | 25.3 (12.7) |
| SCL - Global Severity Index, mean (SD) | 0.98 (0.6) | 0.21 (0.12) | 1.22 (0.6) | 0.79 (0.42) | 1.16 (0.53) | 1.38 (0.58) |
| Family history any psychiatric disorder, N (%) | 284 (60%) | 26 (46%) | 7 (44%) | 96 (64%) | 82 (57%) | 73 (71%) |
| Family History of MDD, N (%) | 207 (43%) | 18 (32%) | 4 (24%) | 73 (48%) | 64 (44%) | 48 (46%) |
| CTQ sum score, mean (SD) | 46.11 (15.7) | 32.4 (5.7) | 50.9 (8.3) | 47.8 (9.8) | 35.8 (7.2) | 64.6 (16.9) |
| Quality of life - mental health, mean (SD) | 48.1 (21.2) | 78.0 (9.1) | 37.4 (19.6) | 53.3 (18.5) | 40.4 (16.8) | 36.9 (17.3) |
| Quality of life - general health, mean (SD) | 52.9 (22.5) | 75.8 (15.3) | 42.9 (26.4) | 60.5 (19.9) | 45.2 (19.5) | 41.9 (19.7) |
| Quality of life – Energy, mean (SD) | 34.4 (20.8) | 61.1 (15.6) | 20.3 (16.4) | 40.1 (17.3) | 26.2 (16.8) | 25.9 (18.5) |
| SCL – Somatization, mean (SD) | 10.8 (8.2) | 3.1 (2.5) | 15.0 (7.6) | 7.0 (4.8) | 14.1 (7.98) | 15.1 (9.1) |
| Life stress, mean (SD) | 29.9 (10.3) | 16.1 (5.9) | 34 (8.7) | 28 (8.8) | 34 (8.7) | 33.8 (9.2) |
| NEO-FFI Neuroticism, mean (SD) | 29.3 (9.0) | 18.1 (6.5) | 33.4 (7.7) | 29.5 (8.1) | 30.8 (8.1) | 32.4 (8.1) |
| Maternal bonding, care, mean (SD) | 20.8 (8.6) | 27.4 (5.5) | 13.9 (5.1) | 18.7 (7.3) | 26.3 (6.2) | 13.7 (7.3) |
| Paternal bonding, care, mean (SD) | 18.7 (8.3) | 24.2 (5.9) | 16.9 (8.8) | 16.6 (7.2) | 21.9 (7.5) | 14.6 (8.6) |
| Positive Symptoms, mean (SD) | 0.79 (2.4) | 0.11 (0.41) | 0.18 (0.52) | 0.41 (1.19) | 0.63 (1.57) | 2.0 (4.4) |
| Antidepressants, yes, N (%) | 292 (61%) | 18 (32%) | 12 (71%) | 84 (55%) | 106 (72%) | 72 (69%) |
| Antipsychotic, yes, N (%) | 84 (18%) | 4 (7%) | 2 (12%) | 26 (17%) | 28 (19%) | 24 (23%) |
| Mood stabilizer, yes, N (%) | 18 (4%) | 2 (4%) | 1 (6%) | 1 (0.7%) | 5 (4%) | 9 (9%) |
| Significantly different PGSs (*one-vs-all*) | | | | | | |
| EA | *p*-value W-Y adjusted | | | | | $1 \times 10^{-3}$ |
| | Bonferroni-corrected (N=5) | | | | | $5 \times 10^{-3}$ |
| ADHD (*) | *p*-value W-Y adjusted | | | | | $1.9 \times 10^{-3}$ |
| | Bonferroni-corrected (N=5) | | | | | $9.5 \times 10^{-3}$ |

**Table S7.** *One-vs-all* HDDA classification analysis using only MDD-diagnosed patients

This table shows the most important clinical variables for each cluster when analyzing only MDD patients. The variables were identified by the *one-vs.-all* HDDA classification analysis using n=477 MDD patients and their respective cluster labels. The importance was calculated based on the average AUC drop, as explained in the Methods S5.

| Cluster 0 | Cluster 1 | Cluster 2 | Cluster 3 | Cluster 4 |
|---|---|---|---|---|
| CTQ Sexual abuse | Maternal bonding, care | SF36 Physical functioning | Maternal bonding, care | CTQ Physical abuse |
| SAPS (Positive symptoms) | SF36 Bodily pain | SF36 Role physical | CTQ Emotional neglect | CTQ Sexual abuse |
| CTQ Physical neglect | SF36 Role physical | CTQ Physical abuse | CTQ Emotional abuse | SAPS (Positive Symptoms) |
| LEQ Negative Events score | YMRS | CTQ Emotional neglect | SF36 Role Physical | CTQ Emotional abuse |
| CTQ Physical abuse | SF36 Energy | RSQ Avoidance of closeness | CTQ Physical abuse | ACE Sum |
| ACE Sum | SHAPS | CTQ Emotional abuse | CTQ Physical neglect | LEQ Positive Events score |
| SCL Positive symptoms | Social support | ACE Sum | ACE Sum | SANS (Negative symptoms) |
| SF36 Social functioning | Verbal IQ | Maternal bonding, care | SF36 Bodily pain | SCL90R Phobic fear |
| SANS (Negative symptoms) | SAPS (Positive symptoms) | CTQ Sexual abuse | SF36 General health | VLMT Sum |
| SF36 Mental health | CTQ Emotional neglect | SCL90R Somatization | SF36 Role emotional | CTQ Physical neglect |

**Table S8.** Metrics of genetic lasso regularized regression prediction models

The table shows the results of lasso regularized regression models applied to characterize clusters using genetic variables. The model consisted of 24 predictors – ten PGS, four family history variables, eight ancestry components, age, and gender, and was applied both in *one-vs-all* and *one-vs-one* analyses. In the one-*vs.*-one comparisons, the cluster tagged as control [27], used for calculation of the sensitivity and specificity, was the respective cluster with the higher rank number. The calculation of all metrics is explained in Methods S5. Only comparisons with an AUC >60% are shown.

| Model | AUC | Sensitivity | Specificity |
|---|---|---|---|
| Cluster 0 *vs.* all | 71% | 66% | 66% |
| Cluster 4 *vs.* all | 73% | 67% | 67% |
| Cluster 0 *vs.* Cluster 4 | 81% | 75% | 75% |
| Cluster 0 *vs.* Cluster 2 | 67% | 67% | 63% |
| Cluster 2 *vs.* Cluster 4 | 67% | 64% | 62% |
| Cluster 0 *vs.* Cluster 3 | 66% | 63% | 63% |
| Cluster 3 *vs.* Cluster 4 | 64% | 61% | 60% |

**Table S9.** Significance testing of genetic analyses with the Westfall and Young procedure – *one-vs-all* comparisons

This table shows the results of *one-vs-all* significance testing of genetic analyses with the Westfall and Young procedure (Methods S6). As in all genetic analyses, 24 variables (ten PGS, four family history variables, eight ancestry components, age, and gender) were used to test for significant differences. Only the comparisons with significant variables after the Westfall and Young adjustment are shown.
The distribution of significant PGS per cluster is shown in Figure 1 E-H.
n.s.: adjusted $p$>0.05; Any = any assessed psychiatric disorder (MDD, BD, SCZ/SZA).

| One-*vs*-All Comparison | Variable | *p*-value adjusted using Westfall and Young | *p*-value further adjusted for the number of comparisons (N=5) |
|---|---|---|---|
| Cluster 0 *vs.* all | Age | $8 \times 10^{-4}$ | $4 \times 10^{-3}$ |
| | Family History Any | $8 \times 10^{-4}$ | $4 \times 10^{-3}$ |
| | Family History BD | $8 \times 10^{-4}$ | $4 \times 10^{-3}$ |
| | Family History MDD | $8 \times 10^{-4}$ | $4 \times 10^{-3}$ |
| | PGS Cross psychiatric disorder | $8 \times 10^{-4}$ | $4 \times 10^{-3}$ |
| | PGS MDD | $1 \times 10^{-3}$ | $8 \times 10^{-3}$ |
| | PGS Schizophrenia | $9 \times 10^{-3}$ | $4 \times 10^{-2}$ |
| | PGS Educational attainment | $2 \times 10^{-2}$ | $9 \times 10^{-2}$ (n.s.) |
| | PGS Neuroticism | $3 \times 10^{-2}$ | $1 \times 10^{-1}$ (n.s.) |
| Cluster 2 *vs.* all | Family History Any | $1 \times 10^{-3}$ | $5 \times 10^{-3}$ |
| | Family History MDD | $6 \times 10^{-3}$ | $3 \times 10^{-2}$ |
| Cluster 4 *vs.* all | Age | $9 \times 10^{-4}$ | $4 \times 10^{-3}$ |
| | Family History Any | $9 \times 10^{-4}$ | $4 \times 10^{-3}$ |
| | PGS Cross psychiatric disorder | $3 \times 10^{-3}$ | $1 \times 10^{-2}$ |
| | PGS MDD | $7 \times 10^{-3}$ | $4 \times 10^{-2}$ |
| | PGS Schizophrenia | $3 \times 10^{-3}$ | $1 \times 10^{-2}$ |
| | PGS Educational attainment | $9 \times 10^{-4}$ | $4 \times 10^{-3}$ |

**Table S10.** Significance testing of genetic analyses with the Westfall and Young procedure – *one-vs-one* comparisons

This table shows the results of *one-vs-one* significance testing of genetic analyses with the Westfall and Young procedure (Methods S6). As in all genetic analyses, 24 variables (ten PGS, four family history variables, eight ancestry components, age, and gender) were used to test for significant differences. Only the comparisons with significant variables after the Westfall and Young adjustment are shown. The distribution of significant PGS per cluster is shown in Figure 1 E-H.
n.s.: adjusted $p$>0.05; Any = any assessed psychiatric disorder (MDD, BD, SCZ/SZA).

| One-*vs*-One Comparison | Variable | $p$-value adjusted using Westfall and Young | $p$-value further adjusted for the number of comparisons (N=10) |
|---|---|---|---|
| Cluster 0 *vs.* Cluster 2 | Age | $1 \times 10^{-3}$ | $1 \times 10^{-2}$ |
| | Family History Any | $1 \times 10^{-3}$ | $1 \times 10^{-2}$ |
| | Family History MDD | $1 \times 10^{-3}$ | $1 \times 10^{-2}$ |
| | PGS Cross psychiatric disorder | $8 \times 10^{-3}$ | $8 \times 10^{-2}$ (n.s.) |
| | PGS MDD | $1 \times 10^{-2}$ | $1 \times 10^{-1}$ (n.s.) |
| | PGS Neuroticism | $1 \times 10^{-2}$ | $1 \times 10^{-1}$ (n.s.) |
| Cluster 0 *vs.* Cluster 3 | Age | $1 \times 10^{-3}$ | $1 \times 10^{-2}$ |
| | Family History Any | $1 \times 10^{-3}$ | $1 \times 10^{-2}$ |
| | Family History MDD | $1 \times 10^{-3}$ | $1 \times 10^{-2}$ |
| Cluster 0 *vs.* Cluster 4 | Age | $8 \times 10^{-4}$ | $8 \times 10^{-3}$ |
| | AC4 | $3 \times 10^{-2}$ | $3 \times 10^{-1}$ (n.s.) |
| | Family History Any | $8 \times 10^{-4}$ | $8 \times 10^{-3}$ |
| | Family History BD | $1 \times 10^{-2}$ | $1 \times 10^{-1}$ (n.s.) |
| | Family History MDD | $1 \times 10^{-3}$ | $1 \times 10^{-2}$ |
| | PGS Cross psychiatric disorder | $8 \times 10^{-4}$ | $8 \times 10^{-3}$ |
| | PGS Schizophrenia | $8 \times 10^{-4}$ | $8 \times 10^{-3}$ |
| | PGS Educational attainment | $8 \times 10^{-4}$ | $8 \times 10^{-3}$ |
| Cluster 2 *vs.* Cluster 4 | Age | $4 \times 10^{-3}$ | $4 \times 10^{-2}$ |
| | Family History BD | $2 \times 10^{-2}$ | $2 \times 10^{-1}$ (n.s.) |
| Cluster 3 *vs.* Cluster 4 | Family History BD | $4 \times 10^{-2}$ | $4 \times 10^{-1}$ (n.s.) |
| | PGS Educational attainment | $3 \times 10^{-2}$ | $3 \times 10^{-1}$ (n.s.) |

**Table S11.** Assessment of the PGS information gain

The table shows the results of multinomial regression analysis for the assessment of the information gain by PGS in the prediction of clusters, explained in the Methods S7. The null model for the calculation of the pseudo-$R^2$ contained only the covariates age and gender.
Abbreviations: AIC = Akaike information criterion; Y = outcome variable (cluster labels); PGSs = all ten polygenic scores (see Methods S3); Family History = four self-reported family history variables (any psychiatric disorder, MDD, BD, SCA/SCZ); ACs = eight ancestry components; n.s. = not significant (adjusted $p$>0.05).

| | **Model A** | **Model B** | **Model C** | **Model D** |
|---|---|---|---|---|
| **Regression model** | Y ~ Age + Gender + PGSs + ACs | Y ~ Age + Gender + Family History | Y ~ Age + Gender + Family History + ACs | Y ~ Age + Gender + Family History + PGSs + ACs (*full model*) |
| **AIC** | 3076.2 | 2975.5 | 2999.1 | 2991.4 |
| **Nagelkerke's pseudo-*R*²** | 11.7% | 10.8% | 13.9% | 20.3% |
| **Likelihood ratio test** | Model A *vs.* Model D | Model B v*s.* Model D | Model B *vs.* Model C | Model C *vs.* Model D |
| **Likelihood ratio test *p*-value** | $2\times10^{-17}$ | $5\times10^{-5}$ | 0.1 (*n.s.*) | $2\times10^{-5}$ |

**Table S12.** Significance testing with the Westfall and Young procedure – comparison of discovery-replication pairs

A matching cluster assignment was confirmed for four clusters. n.s.: adjusted $p$>0.05.

| Discovery-Replication cluster pair | Significant variable | *p*-value adjusted using Westfall and Young | *p*-value further adjusted for the number of comparisons (N=5) |
|---|---|---|---|
| Cluster 0 – Cluster 0 | SF36 General health | $2 \times 10^{-3}$ | $1 \times 10^{-1}$ |
| | NEO-FFI Agreeableness | $4 \times 10^{-3}$ | $2 \times 10^{-2}$ |
| | Letter Number Span test | $4 \times 10^{-3}$ | $2 \times 10^{-2}$ |
| | Paternal Bonding | $3 \times 10^{-2}$ | $1 \times 10^{-1}$ (n.s.) |
| | CTQ Physical Neglect | $4 \times 10^{-2}$ | $2 \times 10^{-1}$ (n.s.) |
| Cluster 2 – Cluster 2 | NEO-FFI Extraversion | $4 \times 10^{-3}$ | $2 \times 10^{-2}$ |
| | NEO-FFI Neuroticism | $1 \times 10^{-2}$ | $5.7 \times 10^{-1}$ (n.s.) |
| | CTQ Emotional Neglect | $2 \times 10^{-2}$ | $8 \times 10^{-2}$ (n.s.) |
| | RS25 Sum Score | $2 \times 10^{-2}$ | $8 \times 10^{-2}$ (n.s.) |
| | RSQ Avoidance of closeness | $4 \times 10^{-2}$ | $2 \times 10^{-1}$ (n.s.) |
| | RSQ Lack of trust | $4 \times 10^{-2}$ | $2 \times 10^{-1}$ (n.s.) |
| Cluster 3 – Cluster 3 | SCL90R Paranoid ideation | $4 \times 10^{-3}$ | $2 \times 10^{-2}$ |
| | SCL90R Somatization | $6 \times 10^{-3}$ | $3 \times 10^{-2}$ |
| | SCL90R Psychoticism | $6 \times 10^{-3}$ | $3 \times 10^{-2}$ |
| | SCL90R Global severity index | $1 \times 10^{-2}$ | $8 \times 10^{-2}$ (n.s.) |
| | SF36 Physical functioning | $2 \times 10^{-2}$ | $1 \times 10^{-1}$ (n.s.) |
| | SCL90R Interpersonal sensitivity | $4 \times 10^{-2}$ | $2 \times 10^{-1}$ (n.s.) |
| Cluster 4 – Cluster 4 | CTQ Physical Neglect | $7 \times 10^{-3}$ | $3 \times 10^{-2}$ |
| | SAPS | $9 \times 10^{-3}$ | $4 \times 10^{-2}$ |
| | SANS | $1.5 \times 10^{-2}$ | $7 \times 10^{-2}$ (n.s.) |

**Table S13.** Characterization of the replication sample and its clusters with variables used in the clustering process

| Category | Variable, mean (SD) | Full | Cluster 0 | Cluster 1 | Cluster 2 | Cluster 3 | Cluster 4 |
|---|---|---|---|---|---|---|---|
| | N | 622 | 128 | 178 | 59 | 136 | 121 |
| Attachment style | RSQ Anxiety of separation | 2.7 (0.7) | 2.4 (0.5) | 2.5 (0.6) | 3.2 (0.8) | 2.7 (0.8) | 2.95 (0.9) |
| | RSQ Avoidance of closeness | 2.5 (0.9) | 1.9 (0.6) | 2.4 (0.7) | 3.3 (0.8) | 2.4 (0.8) | 2.96 (0.9) |
| | RSQ Desire for independence | 3.97 (0.7) | 3.9 (0.6) | 3.95 (0.7) | 3.96 (0.7) | 3.9 (0.8) | 4.1 (0.7) |
| | RSQ Lack of trust | 2.4 (0.9) | 1.7 (0.5) | 2.3 (0.6) | 3.1 (0.8) | 2.3 (0.8) | 3 (0.9) |
| Depression and anxiety level | BDI-II Sum | 11.3 (10.3) | 2.9 (2.8) | 6.6 (5.7) | 14.6 (8.4) | 15.8 (9.1) | 20.4 (12.1) |
| | HAMA Sum | 8.4 (8.2) | 2.5 (3.0) | 4.8 (4.5) | 9.7 (6.8) | 11.6 (7.5) | 15.7 (10.2) |
| | HAMD Sum21 | 6.1 (6.8) | 1.3 (2.1) | 2.9 (3.2) | 7.4 (6.4) | 8.8 (6.5) | 12.2 (7.9) |
| | STAIS | 43 (12.8) | 32.5 (6.0) | 37.3 (8.8) | 48.0 (9.4) | 50.7 (11.8) | 51.6 (13.1) |
| | STAIT | 44.2 (13.8) | 31.3 (6.5) | 38.3 (9.9) | 52.8 (9.8) | 51.8 (11.3) | 53.9 (13.4) |
| Anhedonia | SHAPS | 1.97 (2.7) | 0.4 (0.8) | 0.9 (1.4) | 3.0 (2.4) | 3.3 (3.2) | 3.2 (3.4) |
| Life events | LEQ Negative Events score | 10.4 (12.3) | 3.1 (3.4) | 6.1 (5.8) | 11.2 (8.9) | 11.8 (8.6) | 22.6 (19.1) |
| | LEQ Positive Events score | 9.4 (9.3) | 8.2 (7.1) | 9.6 (7.9) | 10.2 (8.3) | 7.2 (7.5) | 12.7 (13.8) |
| Life stress | PSS Sum | 23.8 (10.5) | 14.4 (6.2) | 19.3 (7.4) | 27.9 (7.1) | 29.5 (8.7) | 31.9 (10.5) |
| Maltreatment in childhood and youth | ACE Sum | 1.6 (1.8) | 0.4 (0.6) | 1.3 (1.3) | 2.9 (1.6) | 1.1 (1.3) | 3.1 (2.3) |
| | CTQ Emotional abuse | 9.1 (4.5) | 6.1 (1.5) | 8.2 (2.8) | 13.4 (4.8) | 7.8 (3.0) | 13.2 (5.5) |
| | CTQ Emotional neglect | 11.1 (5.2) | 6.9 (2.1) | 10.8 (3.7) | 16.6 (4.7) | 9.4 (3.4) | 14.98 (6.1) |
| | CTQ Physical abuse | 6.1 (2.4) | 5.2 (0.6) | 5.7 (1.3) | 6.0 (1.7) | 5.5 (1.0) | 8.4 (4.1) |
| | CTQ Physical neglect | 6.8 (2.5) | 5.4 (1.0) | 6.5 (1.4) | 8.5 (2.3) | 6.1 (1.7) | 8.8 (3.6) |
| | CTQ Sexual abuse | 5.7 (2.5) | 5 (0.3) | 5.3 (1.0) | 5.3 (1.0) | 5.3 (1.2) | 7.8 (4.8) |
| Mania Symptoms | YMRS | 1.4 (2.7) | 0.3 (0.7) | 1.2 (1.7) | 1.3 (2.2) | 1.3 (1.7) | 3.2 (4.6) |
| Negative symptoms | SANS sum score | 4.3 (7.3) | 0.3 (0.9) | 1.7 (3.1) | 3.96 (5.6) | 6.4 (7.3) | 10.4 (10.7) |
| Positive symptoms | SAPS sum score | 0.7 (2.7) | 0.04 (0.2) | 0.2 (0.6) | 0.5 (1.4) | 0.4 (1.2) | 2.8 (5.3) |

| Category | Variable | Full | Cluster 0 | Cluster 1 | Cluster 2 | Cluster 3 | Cluster 4 |
|---|---|---|---|---|---|---|---|
| Personality | NEO-FFI Agreeableness | 33.2 (6.1) | 37.4 (4.1) | 32.6 (5.3) | 30.0 (5.7) | 34.5 (5.6) | 29.96 (6.6) |
| | NEO-FFI Conscientiousness | 32.2 (7.4) | 36.3 (6) | 32.7 (6.4) | 29.5 (6.9) | 30.9 (7.4) | 29.9 (8.2) |
| | NEO-FFI Extraversion | 25.8 (8.2) | 32 (5.8) | 26.7 (6.8) | 19.6 (6.3) | 24.1 (8.2) | 22.7 (8.5) |
| | NEO-FFI Neuroticism | 23.1 (10.2) | 13.9 (6.6) | 19.5 (8.2) | 30.4 (7.3) | 26.7 (8.7) | 30.7 (8.6) |
| | NEO-FFI Openness to experience | 30.4 (6.8) | 32.7 (5.4) | 30.7 (6.2) | 29.5 (7.6) | 29.2 (6.8) | 29.3 (8) |
| | SPQB (Schizotypy) | 5.97 (4.7) | 2.4 (2.4) | 5.1 (3.6) | 9.6 (4.9) | 5.8 (4.2) | 9.4 (4.8) |
| Protective factors | Maternal bonding, care | 24.4 (8.5) | 30.9 (4.1) | 24.1 (6.6) | 17.4 (8.6) | 26.8 (6.9) | 18.4 (9.6) |
| | Paternal bonding, care | 21.2 (8.9) | 28.8 (5) | 19.7 (7.2) | 14.1 (7.9) | 23.9 (7.7) | 16.1 (9.2) |
| | RS25 Sum Score (Resilience) | 125.1 (26.9) | 145.1 (14.3) | 134.0 (22.0) | 105.3 (22.6) | 116.9 (24.7) | 109.8 (28.8) |
| | Social support | 4.1 (0.7) | 4.7 (0.3) | 4.3 (0.5) | 3.5 (0.8) | 4.1 (0.7) | 3.7 (0.9) |
| Short Form Health Survey / Quality of life | SF36 Bodily pain | 74.7 (26.1) | 90.5 (13.3) | 83.1 (18.6) | 77.3 (21.7) | 66.3 (28.9) | 53.9 (27.9) |
| | SF36 Energy/fatigue | 47.5 (22.1) | 66.4 (13.1) | 58.1 (16.6) | 38.98 (15.3) | 30.8 (15.1) | 34.6 (22.5) |
| | SF36 General health | 68.1 (23) | 88.6 (12.8) | 75.9 (16.3) | 64.5 (16.6) | 56.3 (20.8) | 49.8 (22.4) |
| | SF36 Mental health | 62.8 (22.1) | 83.7 (9.4) | 73.4 (13.8) | 55.1 (16.8) | 48.2 (17.2) | 45.5 (22) |
| | SF36 Physical functioning | 89.8 (15.5) | 97.7 (4.6) | 94.8 (8.6) | 92.5 (8.9) | 85.6 (16.5) | 77.4 (21.9) |
| | SF36 Role emotional | 60.6 (43.2) | 95.8 (13.2) | 81.1 (31.1) | 47.5 (40.7) | 27.9 (36.3) | 36.4 (44.1) |
| | SF36 Role physical | 71.2 (36.1) | 97.1 (10.2) | 89.2 (17.8) | 70.8 (28.7) | 47.1 (37) | 44.6 (40.9) |
| | SF36 Social functioning | 70.4 (30.1) | 95.7 (9.1) | 85.6 (17.6) | 63.8 (28.3) | 50.9 (26.5) | 46.3 (30.1) |
| Symptom checklist | SCL90R Additional Items | 4.4 (3.8) | 1.6 (1.6) | 2.8 (2.1) | 5.6 (3.8) | 5.6 (3.4) | 7.5 (4.6) |
| | SCL90R Anxiety | 5.4 (6.2) | 1.1 (1.4) | 2.6 (2.5) | 6 (5.6) | 7.4 (5.6) | 11.4 (8.1) |
| | SCL90R Depression | 12.2 (11.6) | 2.7 (3.3) | 6.5 (5.8) | 16.1 (10.1) | 18.0 (9.8) | 22.4 (13.4) |
| | SCL90R Global severity index | 0.6 (0.5) | 0.2 (0.1) | 0.3 (0.2) | 0.8 (0.4) | 0.8 (0.4) | 1.2 (0.7) |
| | SCL90R Hostility | 3.2 (3.9) | 0.9 (1.2) | 1.7 (1.8) | 4.3 (3.4) | 3.9 (3.5) | 6.6 (5.5) |
| | SCL90R Interpersonal sensitivity | 6.96 (7.0) | 1.7 (2.1) | 4.1 (3.6) | 11.2 (6.8) | 8.0 (5.6) | 13.6 (8.8) |

| Category | Variable | Full | Cluster 0 | Cluster 1 | Cluster 2 | Cluster 3 | Cluster 4 |
|---|---|---|---|---|---|---|---|
| Symptom checklist | SCL90R Obsessive-compulsive behavior | 8.7 (7.7) | 2.5 (2.6) | 4.7 (3.99) | 11.3 (5.9) | 12.7 (6.5) | 15.1 (9.0) |
| | SCL90R Paranoid ideation | 3.3 (4.0) | 0.7 (1.1) | 1.9 (2.0) | 4.7 (3.5) | 3.3 (3.0) | 7.3 (5.7) |
| | SCL90R Phobic anxiety | 2.3 (3.9) | 0.2 (0.7) | 0.6 (1.1) | 2.5 (2.7) | 2.8 (3.1) | 6.3 (6.2) |
| | SCL90R Positive symptom distress Index | 1.5 (0.5) | 1.1 (0.2) | 1.2 (0.2) | 1.6 (0.4) | 1.7 (0.4) | 2.0 (0.6) |
| | SCL90R Positive symptom total | 31.2 (19.5) | 12.5 (9.1) | 22.7 (11.9) | 41.7 (15.9) | 40.0 (14.5) | 48.4 (19.9) |
| | SCL90R Psychoticism | 3.7 (4.9) | 0.5 (1.0) | 1.6 (1.8) | 5.7 (4.1) | 4.4 (3.9) | 8.6 (7.1) |
| | SCL90R Somatization | 7.1 (6.5) | 2.8 (2.1) | 4.2 (3.2) | 8.0 (5.9) | 9.3 (5.7) | 13.0 (8.7) |
| Verbal IQ | Verbal IQ | 113.8 (13.4) | 117.4 (13.4) | 116.4 (13.8) | 110.1 (12.4) | 112.5 (12.2) | 109.3 (12.9) |
| Verbal Learning and memory test | VLMT Sum | 56.1 (9.6) | 60.6 (6.8) | 56.2 (9.5) | 54.7 (8.9) | 55.6 (9.6) | 52.4 (10.9) |
| Visuospatial Working Memory Test | Corsi block-tapping test | 17.2 (3.1) | 18.6 (2.6) | 17.5 (2.7) | 17.0 (3.1) | 16.9 (3.4) | 15.6 (3.1) |
| Visuospatial Working Memory Test | Letter Number Span test | 16.1 (3.4) | 17.9 (2.6) | 16.5 (3.2) | 15.7 (3.1) | 15.9 (3.3) | 14.2 (3.6) |

**Table S14.** Projection of the discovery-stage lasso models to the replication sample

Lasso models from the discovery-stage analysis were projected to their matched replication pairs, as explained in the Methods S8.

| **Model projection to the corresponding cluster** | **AUC** | **Sensitivity** | **Specificity** |
|---|---|---|---|
| Cluster 0 *vs.* all | 63% | 60% | 60% |
| Cluster 4 *vs.* all | 68% | 67% | 66% |
| Cluster 0 *vs.* Cluster 4 | 75% | 72% | 72% |
| Cluster 2 *vs.* Cluster 4 | 69% | 72% | 67% |
| Cluster 3 *vs.* Cluster 4 | 61% | 60% | 60% |
| Cluster 0 *vs.* Cluster 2 | 60% | 59% | 59% |
| Cluster 0 *vs.* Cluster 3 | 60% | 70% | 53% |

**Table S15.** Replication – Significance testing with the WY procedure, *one-vs-all* comparisons

Only the comparisons with significant variables after the first adjustment are shown.
n.s.: not significant at an α=0.05 threshold. WY = Westfall and Young.

| One-*vs*-All Comparison | Variable | *p*-value adjusted using Westfall and Young | *p*-value further adjusted for the number of comparisons (N=5) |
|---|---|---|---|
| Cluster 0 *vs.* all | Age | $3 \times 10^{-2}$ | $1 \times 10^{-1}$ (n.s.) |
| | PGS Cross psychiatric disorder | $7 \times 10^{-3}$ | $3 \times 10^{-2}$ |
| | PGS Schizophrenia | $1 \times 10^{-3}$ | $5 \times 10^{-3}$ |
| Cluster 2 *vs.* all | Age | $1 \times 10^{-2}$ | $5 \times 10^{-2}$ |
| Cluster 4 *vs.* all | PGS Cross psychiatric disorder | $1.6 \times 10^{-2}$ | $8 \times 10^{-2}$ (n.s.) |
| | PGS MDD | $3 \times 10^{-3}$ | $1 \times 10^{-2}$ |
| | PGS Schizophrenia | $2 \times 10^{-2}$ | $1 \times 10^{-1}$ (n.s.) |
| | PGS Educational attainment | $1 \times 10^{-3}$ | $5 \times 10^{-3}$ |
| | PGS Neuroticism | $6 \times 10^{-3}$ | $3 \times 10^{-2}$ |

**Table S16.** Replication – Significance testing with the WY procedure, *one-vs-one* comparisons

Only the comparisons with significant variables after the first adjustment are shown.
n.s.: not significant at an α=0.05 threshold. WY = Westfall and Young.

| One-*vs*-One Comparison | Variable | *p*-value adjusted using Westfall and Young | *p*-value further adjusted for the number of comparisons (N=5) |
|---|---|---|---|
| Cluster 0 *vs.* Cluster 1 | Age | $5 \times 10^{-3}$ | $5 \times 10^{-2}$ |
| Cluster 0 *vs.* Cluster 4 | PGS Cross psychiatric disorder | $1 \times 10^{-3}$ | $1 \times 10^{-2}$ |
| | PGS MDD | $9 \times 10^{-3}$ | $9 \times 10^{-2}$ (n.s.) |
| | PGS Schizophrenia | $1 \times 10^{-3}$ | $1 \times 10^{-2}$ |
| | PGS Educational attainment | $2 \times 10^{-3}$ | $2 \times 10^{-2}$ |
| Cluster 1 *vs.* Cluster 2 | Age | $2 \times 10^{-3}$ | $2 \times 10^{-2}$ |
| Cluster 1 *vs.* Cluster 4 | PGS MDD | $1 \times 10^{-2}$ | $1 \times 10^{-1}$ (n.s.) |
| | PGS Educational attainment | $2 \times 10^{-3}$ | $2 \times 10^{-2}$ |
| | PGS Neuroticism | $4 \times 10^{-3}$ | $4 \times 10^{-2}$ |
| Cluster 2 *vs.* Cluster 3 | Age | $2 \times 10^{-2}$ | $2 \times 10^{-1}$ (n.s.) |
| Cluster 2 vs. Cluster 4 | Age | $1 \times 10^{-2}$ | $1 \times 10^{-1}$ (n.s.) |
| | PGS Educational attainment | $1 \times 10^{-2}$ | $1 \times 10^{-1}$ (n.s.) |
| Cluster 3 *vs.* Cluster 4 | PGS Educational attainment | $4 \times 10^{-2}$ | $4 \times 10^{-1}$ (n.s.) |

**Table S17.** Bipolar disorder subtypes across clusters.
NOS = not otherwise specified.

| Variable | Full sample | Cluster 0 | Cluster 1 | Cluster 2 | Cluster 3 | Cluster 4 |
|---|---|---|---|---|---|---|
| **Discovery sample** | | | | | | |
| N (BD diagnosis) | 75 | 9 | 1 | 15 | 26 | 24 |
| Bipolar Type I, N (%) | 41 (55%) | 6 (67%) | 0 (0%) | 3 (20%) | 18 (69%) | 14 (58%) |
| Bipolar Type II, N (%) | 33 (44%) | 2 (22%) | 1 (100%) | 12 (80%) | 8 (31%) | 10 (42%) |
| Bipolar Disorder NOS, N (%) | 1 (1%) | 1 (11%) | 0 (0%) | 0 (0%) | 0 (0%) | 0 (0%) |
| **Replication sample** | | | | | | |
| N (BD diagnosis) | 44 | 1 | 9 | 7 | 17 | 10 |
| Bipolar Type I, N (%) | 21 (48%) | 0 (0%) | 5 (56%) | 2 (29%) | 8 (47%) | 6 (60%) |
| Bipolar Type II, N (%) | 23 (52%) | 1 (100%) | 4 (44%) | 5 (71%) | 9 (53%) | 4 (40%) |
| Bipolar Disorder NOS, N (%) | 0 (0%) | 0 (0%) | 0 (0%) | 0 (0%) | 0 (0%) | 0 (0%) |

**Figure S1.** Population substructure (genetic ancestry components).

Post-QC, pre-imputation genotype data were used to calculate ancestry components (for details on the QC, see the Methods S2).

Additional variant filtering steps were: Removal of variants with a MAF <0.05 or HWE *p*-value $<10^{-3}$; removal of variants mapping to the extended major histocompatibility region (chromosome 6, 25-35 Mbp) or to a typical inversion site on chromosome 8 (7-13 Mbp); linkage disequilibrium (LD) pruning (command `--indep-pairwise 200 100 0.2`).

Next, the pairwise identity-by-state (IBS) matrix of all individuals was calculated using the command `--genome` on the filtered genotype data. Multidimensional scaling (MDS) analysis was performed on the IBS matrix using the eigendecomposition-based algorithm in PLINK v1.90.

Abbreviations: HC, healthy control; MDD, major depressive disorder; BD, bipolar disorder; SZA, schizoaffective disorder; SCZ, schizophrenia; OTH, other psychiatric diagnoses.

**A:** Population substructure in the discovery sample.

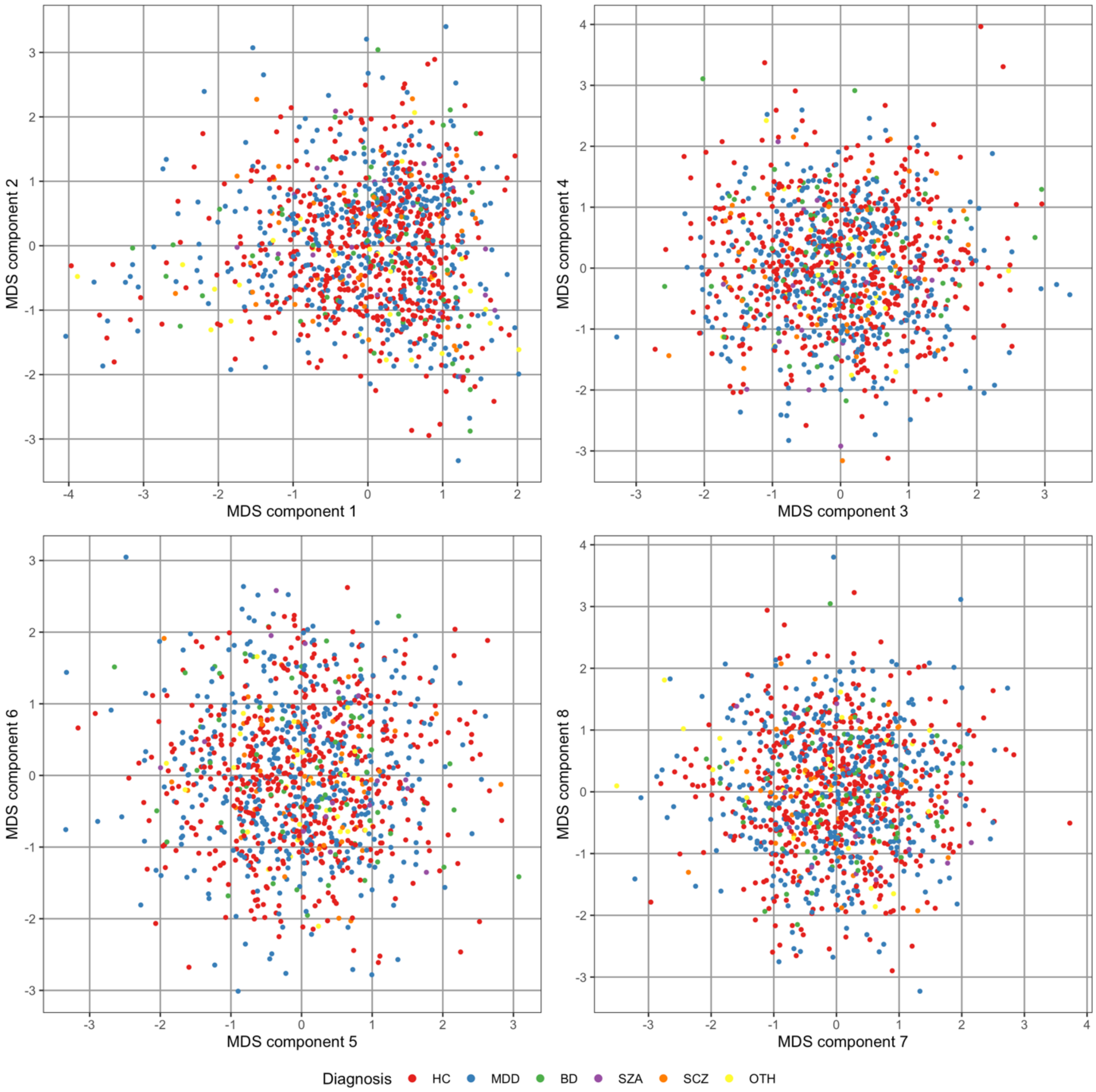

**B:** Population substructure in the replication sample.

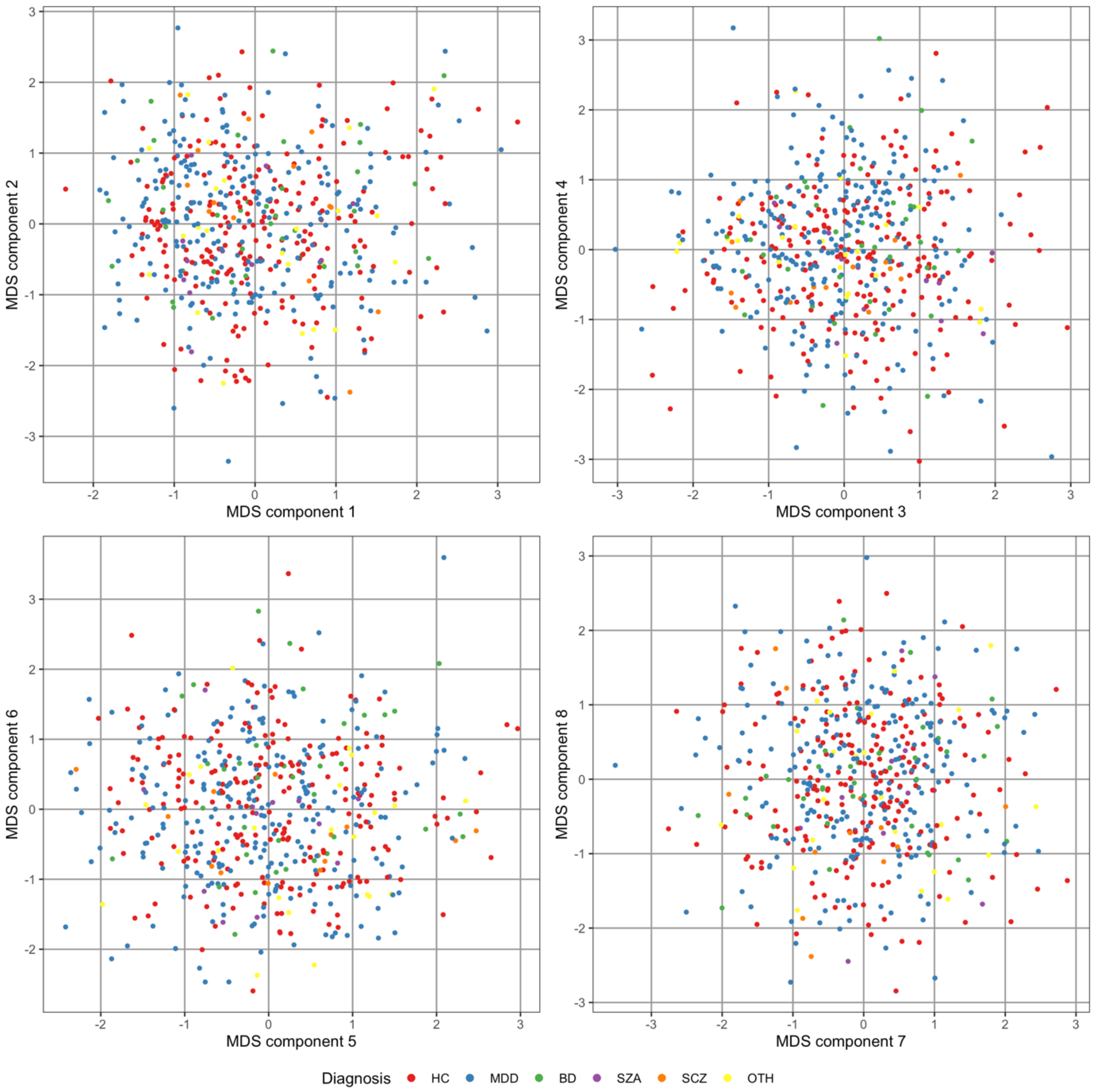

**Figure S2.** Overview of the clustering pipeline
Abbreviations: ICL, Integrated Likelihood criterion [21].

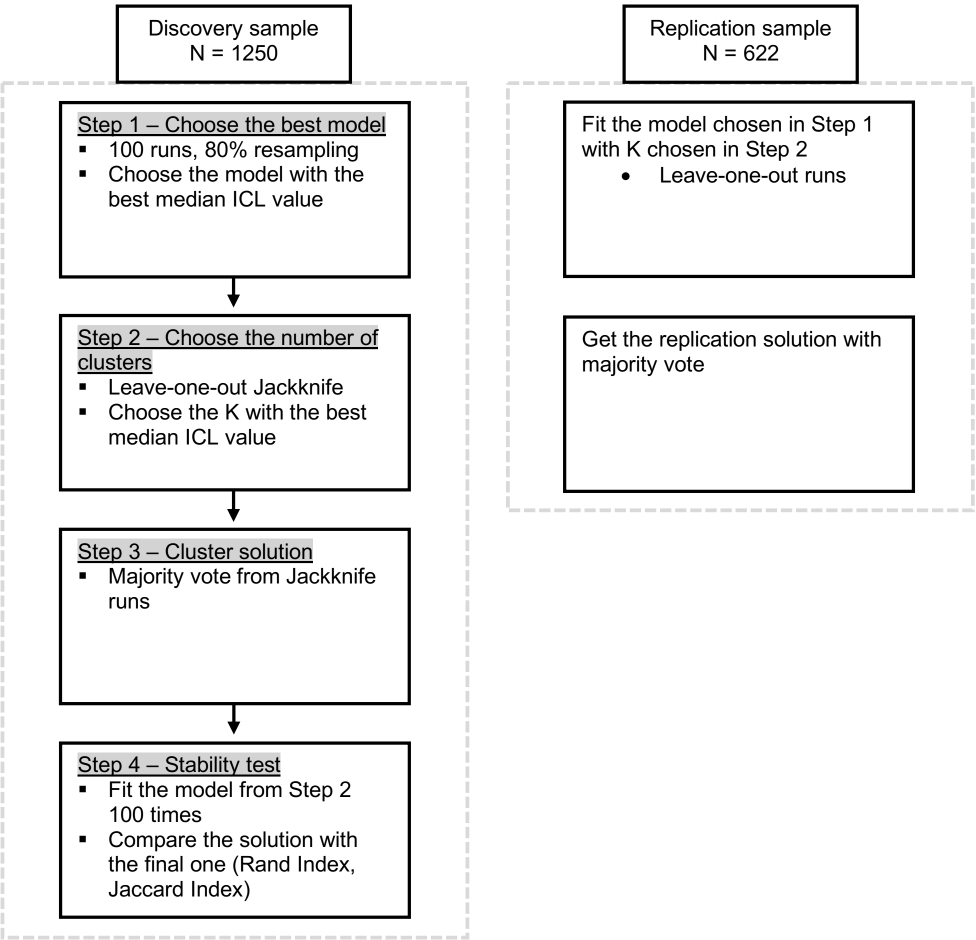

**Figure S3.** Results from the clustering pipeline

**A**: Step 1: Choosing the best fitting model. There were two groups of models created. Models that allow for modeling of the class specific noise are in the better fitting group. Among those, model AkBkQkDk achieved the highest median ICL value and was submitted to the second step. For theoretical details, see Bouveyron *et al.* [19].
**B**: Step 2: Choosing the optimal number of clusters K.
**C:** Stability test: K = 5 confirmed as an optimal solution.
**D:** Stability measures – Rand and Jaccard indices. Stability labels were obtained with the majority voting scheme from the stability analysis runs and compared to the final cluster labels.

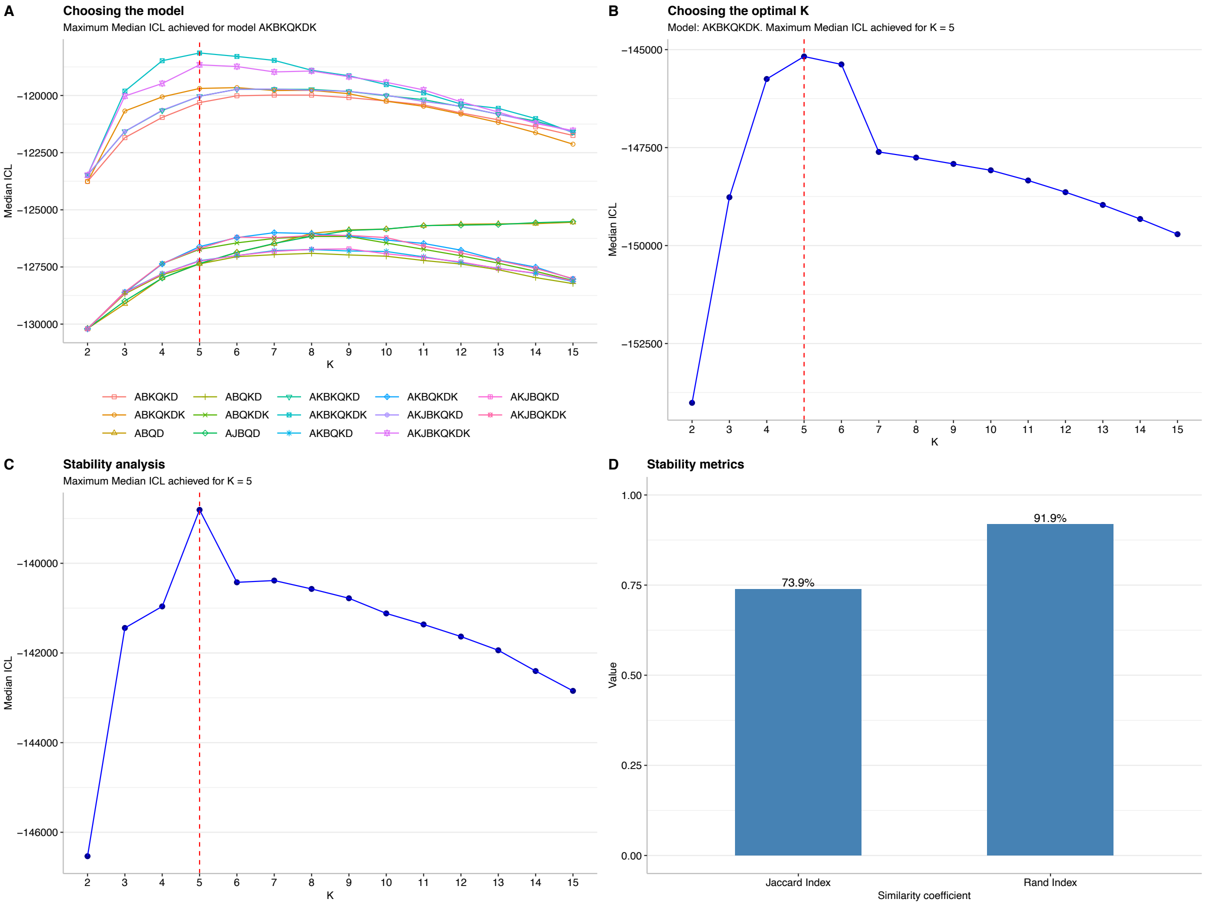

**Figure S4.** Distribution of PGS for MDD patients

The plot shows only PGS that were significant after Bonferroni correction (adjusted *p*<0.05) in *one-vs-all* analyses using the Westfall and Young procedure (Methods S6) **in the discovery-stage analysis of MDD patients** (Table S6). PGS were standardized by Z-score transformation, the y-axis unit is standard deviations. A horizontal line represents the mean and the error bars indicate the standard deviation of the variables in the respective clusters.
Panels A-B show results from the discovery and C-D from the replication sample.

**A:** Attention-deficit/hyperactivity disorder PGS in discovery-stage MDD patients**,** significantly higher for the MDD patients in cluster 4 compared to the MDD patients in all other clusters (adjusted *p*=0.0095). The ADHD PGS was not significant in the analysis including all diagnoses**. B:** Educational attainment PGS in discovery-stage MDD patients, significantly lower for the MDD patients in cluster 4 compared to the MDD patients in all other clusters (adjusted *p*=0.005)**. C**: ADHD PGS for MDD patients in the replication sample, The discovery-stage association (significantly higher ADHD PGS in cluster 4 compared to all other clusters) did not replicate. **D:** PGS Educational attainment for MDD patients in the replication sample. The significant difference of MDD patients in cluster 4 against all other MDD patients replicated (Bonferroni-adjusted *p*=0.005).

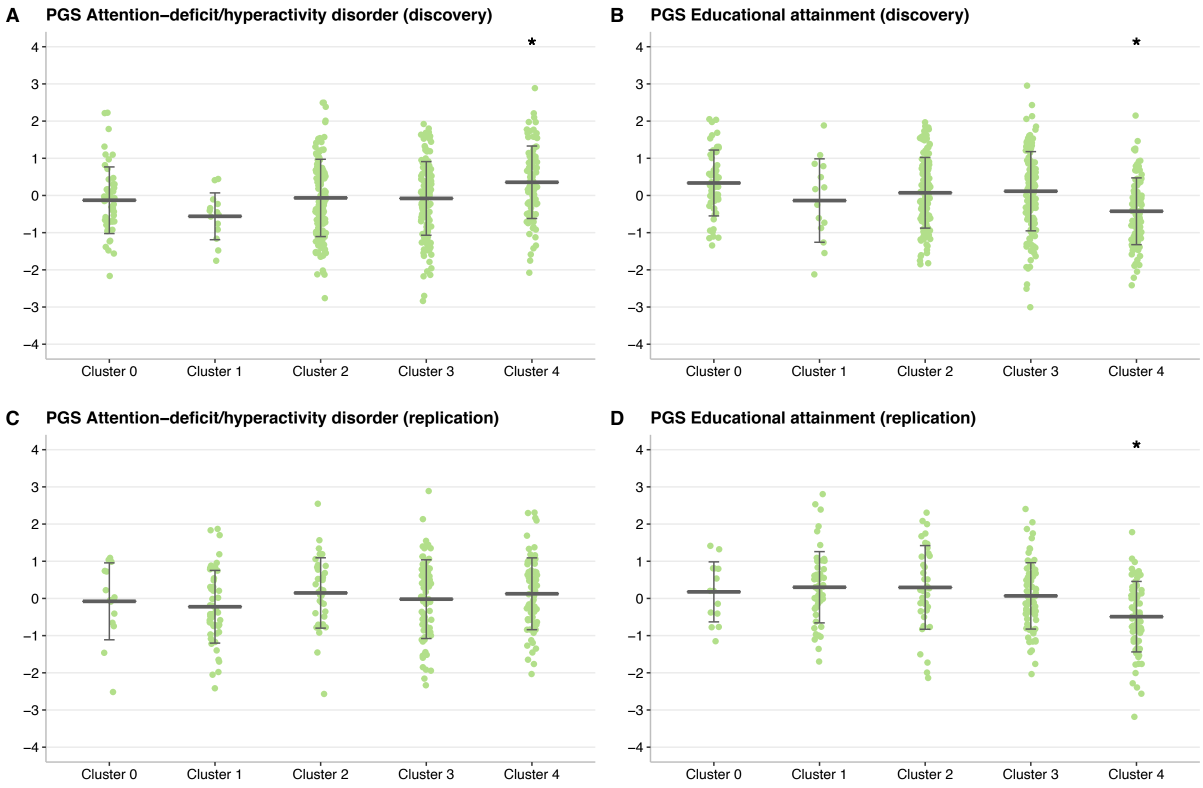

**Figure S5.** Discovery-stage HDDA models projected to replication sample

The projection of HDDA models was done as explained in Methods S8. The matched discovery-stage and replication clusters achieving the prediction>70% are represented with colored bars. All models except *Cluster 1-vs-all* showed higher predictions to the replication sample for one of the projections.

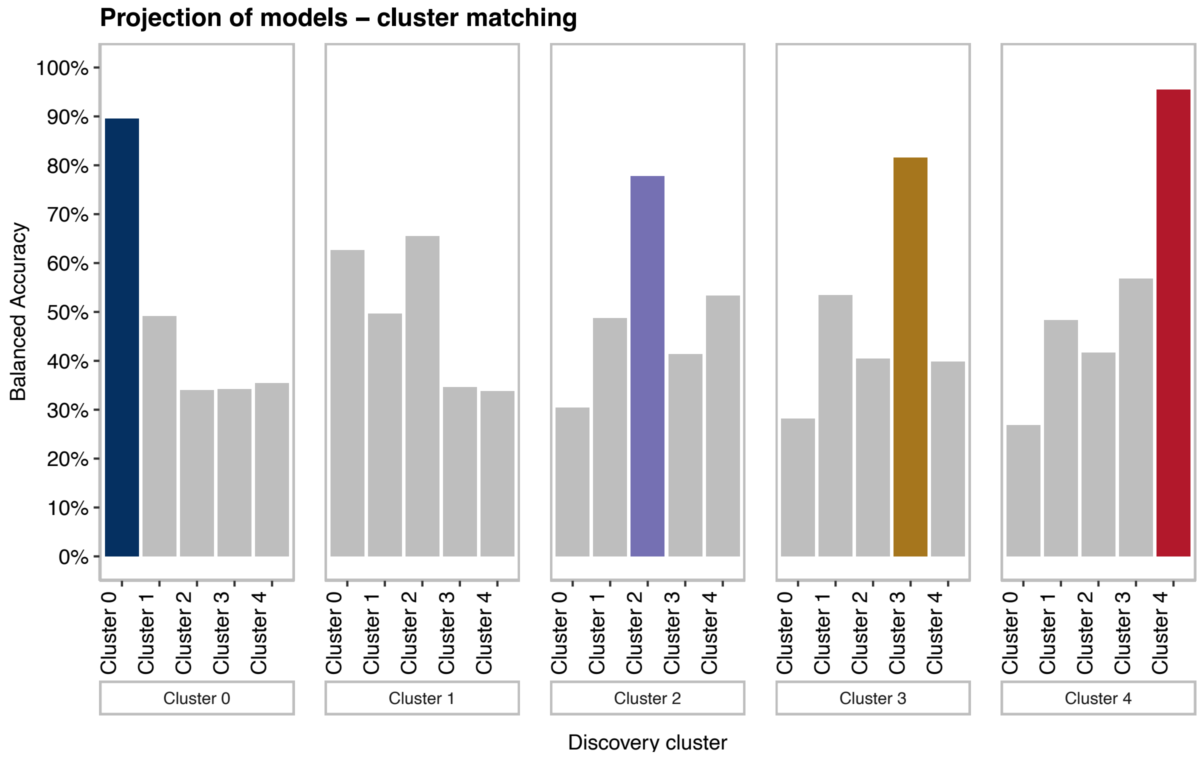

**Full FOR2107 acknowledgements**

This work is part of the German multicenter consortium "Neurobiology of Affective Disorders. A translational perspective on brain structure and function", funded by the German Research Foundation (Deutsche Forschungsgemeinschaft DFG; Forschungsgruppe/Research Unit FOR2107).
Principal investigators (PIs) with respective areas of responsibility in the FOR2107 consortium are: Work Package WP1, FOR2107/MACS cohort and brainimaging: Tilo Kircher (speaker FOR2107; DFG grant numbers KI 588/14-1, KI 588/14-2), Udo Dannlowski (co-speaker FOR2107; DA 1151/5-1, DA 1151/5-2), Axel Krug (KR 3822/5-1, KR 3822/7-2), Igor Nenadic (NE 2254/1-2), Carsten Konrad (KO 4291/3-1). WP2, animal phenotyping: Markus Wöhr (WO 1732/4-1, WO 1732/4-2), Rainer Schwarting (SCHW 559/14-1, SCHW 559/14-2). WP3, miRNA: Gerhard Schratt (SCHR 1136/3-1, 1136/3-2). WP4, immunology, mitochondriae: Judith Alferink (AL 1145/5-2), Carsten Culmsee (CU 43/9-1, CU 43/9-2), Holger Garn (GA 545/5-1, GA 545/7-2). WP5, genetics: Marcella Rietschel (RI 908/11-1, RI 908/11-2), Markus Nöthen (NO 246/10-1, NO 246/10-2), Stephanie Witt (WI 3439/3-1, WI 3439/3-2). WP6, multimethod data analytics: Andreas Jansen (JA 1890/7-1, JA 1890/7-2), Tim Hahn (HA 7070/2-2), Bertram Müller-Myhsok (MU1315/8-2), Astrid Dempfle (DE 1614/3-1, DE 1614/3-2). CP1, biobank: Petra Pfefferle (PF 784/1-1, PF 784/1-2), Harald Renz (RE 737/20-1, 737/20-2). CP2, administration. Tilo Kircher (KI 588/15-1, KI 588/17-1), Udo Dannlowski (DA 1151/6-1), Carsten Konrad (KO 4291/4-1).
Data access and responsibility: All PIs take responsibility for the integrity of the respective study data and their components. All authors and coauthors had full access to all study data.
Acknowledgements and members by Work Package (WP): WP1: Henrike Bröhl, Katharina Brosch, Bruno Dietsche, Rozbeh Elahi, Jennifer Engelen, Sabine Fischer, Jessica Heinen, Svenja Klingel, Felicitas Meier, Tina Meller, Julia-Katharina Pfarr, Kai Ringwald, Torsten Sauder, Simon Schmitt, Frederike Stein, Annette Tittmar, Dilara Yüksel (Dept. of Psychiatry, Marburg University). Mechthild Wallnig, Rita Werner (Core-Facility Brainimaging, Marburg University). Carmen Schade-Brittinger, Maik Hahmann (Coordinating Centre for Clinical Trials, Marburg). Michael Putzke (Psychiatric Hospital, Friedberg). Rolf Speier, Lutz Lenhard (Psychiatric Hospital, Haina). Birgit Köhnlein (Psychiatric Practice, Marburg). Peter Wulf, Jürgen Kleebach, Achim Becker (Psychiatric Hospital Hephata, Schwalmstadt-Treysa). Ruth Bär (Care facility Bischoff, Neukirchen). Matthias Müller, Michael Franz, Siegfried Scharmann, Anja Haag, Kristina Spenner, Ulrich Ohlenschläger (Psychiatric Hospital Vitos, Marburg). Matthias Müller, Michael Franz, Bernd Kundermann (Psychiatric Hospital Vitos, Gießen). Christian Bürger, Katharina Dohm, Fanni Dzvonyar, Verena Enneking, Stella Fingas, Katharina Förster, Janik Goltermann, Dominik Grotegerd, Hannah Lemke, Susanne Meinert, Nils Opel, Ronny Redlich, Jonathan Repple, Kordula Vorspohl, Bettina Walden, Dario Zaremba (Dept. of Psychiatry, University of Münster). Harald Kugel, Jochen Bauer, Walter Heindel, Birgit Vahrenkamp (Dept. of Clinical Radiology, University of Münster). Gereon Heuft, Gudrun Schneider (Dept. of Psychosomatics and Psychotherapy, University of Münster). Thomas Reker (LWL-Hospital Münster). Gisela Bartling (IPP Münster). Ulrike Buhlmann (Dept. of Clinical Psychology, University of Münster). WP5: Helene Dukal, Christine Hohmeyer, Lennard Stütz, Viola Lahr, Fabian Streit, Josef Frank, Lea Sirignano (Dept. of Genetic Epidemiology, Central Institute of Mental Health, Medical Faculty Mannheim, Heidelberg University). Stefanie Heilmann-Heimbach, Stefan Herms, Per Hoffmann (Institute of Human Genetics, University of Bonn, School of Medicine & University Hospital Bonn). Andreas J. Forstner (Institute of Human Genetics, University of Bonn, School of Medicine & University Hospital Bonn; Centre for Human Genetics, Marburg University). WP6: Anastasia Benedyk, Miriam Bopp, Roman Keßler, Maximilian Lückel, Verena Schuster, Christoph Vogelbacher (Dept. of Psychiatry, Marburg University). Jens Sommer, Olaf Steinsträter (Core-Facility Brainimaging, Marburg University). Thomas W.D. Möbius (Institute of Medical Informatics and Statistics, Kiel University). CP1: Julian Glandorf, Fabian Kormann, Arif Alkan, Fatana Wedi, Lea Henning, Alena Renker, Karina Schneider, Elisabeth Folwarczny, Dana Stenzel, Kai Wenk, Felix Picard, Alexandra Fischer, Sandra Blumenau, Beate Kleb, Doris Finholdt, Elisabeth Kinder, Tamara Wüst, Elvira Przypadlo, Corinna Brehm (Comprehensive Biomaterial Bank Marburg, Marburg University).
The FOR2107 cohort project (WP1) was approved by the Ethics Committees of the Medical Faculties, University of Marburg (AZ: 07/14) and University of Münster (AZ: 2014-422-b-S). The study was supported by the German Federal Ministry of Education and Research (BMBF), through the Integrated Network IntegraMent, under the auspices of the e:Med programme (grants 01ZX1314A/01ZX1614A to MMN; 01ZX1314G/01ZX1614G to MR; 01ZX1614J to BMM) through grants 01EE1406C to MR and 01EE1409C to MR and SHW, and through ERA-NET NEURON, "SynSchiz - Linking synaptic dysfunction to disease mechanisms in schizophrenia - a multilevel investigation" (01EW1810 to MR) and BMBF grants 01EE1409C and 01EE1406C to MR and SHW.

**Supplemental Material References**


1. Meller T, Schmitt S, Stein F, Brosch K, Mosebach J, Yüksel D, et al. Associations of schizophrenia risk genes ZNF804A and CACNA1C with schizotypy and modulation of attention in healthy subjects. Schizophr Res. 2019. 7 May 2019.
2. Chang CC, Chow CC, Tellier LCAM, Vattikuti S, Purcell SM, Lee JJ. Second-generation PLINK: rising to the challenge of larger and richer datasets. Gigascience. 2015;4.
3. Andlauer TFM, Buck D, Antony G, Bayas A, Bechmann L, Berthele A, et al. Novel multiple sclerosis susceptibility loci implicated in epigenetic regulation. Sci Adv. 2016;2:e1501678–e1501678.
4. Delaneau O, Zagury J-F, Marchini J. Improved whole-chromosome phasing for disease and population genetic studies. Nat Methods. 2013;10:5–6.
5. Howie B, Fuchsberger C, Stephens M, Marchini J, Abecasis GR. Fast and accurate genotype imputation in genome-wide association studies through pre-phasing. Nat Genet. 2012;44:955–959.
6. Howie BN, Donnelly P, Marchini J. A flexible and accurate genotype imputation method for the next generation of genome-wide association studies. PLoS Genet. 2009;5:e1000529–e1000529.
7. Demontis D, Walters RK, Martin J, Mattheisen M, Als TD, Agerbo E, et al. Discovery of the first genome-wide significant risk loci for attention deficit/hyperactivity disorder. Nat Genet. 2019;51:63–75.
8. Grove J, Ripke S, Als TD, Mattheisen M, Walters RK, Won H, et al. Identification of common genetic risk variants for autism spectrum disorder. Nat Genet. 2019;51:431–444.
9. Stahl EA, Breen G, Forstner AJ, McQuillin A, Ripke S, Trubetskoy V, et al. Genome-wide association study identifies 30 loci associated with bipolar disorder. Nat Genet. 2019;51:793–803.
10. Lee P, Anttila V, Won H, Feng Y-C, Rosenthal J, Zhu Z, et al. Genomic Relationships, Novel Loci, and Pleiotropic Mechanisms across Eight Psychiatric Disorders. Cell. 2019;179:1469–1482.
11. Okbay A, Beauchamp JP, Fontana MA, Lee JJ, Pers TH, Rietveld CA, et al. Genome-wide association study identifies 74 loci associated with educational attainment. Nature. 2016;533:539–542.
12. van den Berg SM, de Moor MHM, Verweij KJH, Krueger RF, Luciano M, Arias Vasquez A, et al. Meta-analysis of Genome-Wide Association Studies for Extraversion: Findings from the Genetics of Personality Consortium. Behav Genet. 2016;46:170–182.
13. Baselmans BML, Bartels M. A genetic perspective on the relationship between eudaimonic -and hedonic well-being. Sci Rep. 2018;8:14610.
14. Howard DM, Adams MJ, Clarke T-K, Hafferty JD, Gibson J, Shirali M, et al. Genome-wide meta-analysis of depression identifies 102 independent variants and highlights the importance of the prefrontal brain regions. Nat Neurosci. 2019;22:343–352.
15. Luciano M, Hagenaars SP, Davies G, Hill WD, Clarke T-K, Shirali M, et al. Association analysis in over 329,000 individuals identifies 116 independent variants influencing neuroticism. Nat Genet. 2018;50:6–11.
16. Pardiñas AF, Holmans P, Pocklington AJ, Escott-Price V, Ripke S, Carrera N, et al. Common schizophrenia alleles are enriched in mutation-intolerant genes and in regions under strong background selection. Nat Genet. 2018;50:381–389.
17. Ge T, Chen C-Y, Ni Y, Feng Y-CA, Smoller JW. Polygenic prediction via Bayesian regression and continuous shrinkage priors. Nat Commun. 2019;10:1776.
18. Andlauer TFM, Guzman-Parra J, Streit F, Strohmaier J, González MJ, Gil Flores S, et al. Bipolar multiplex families have an increased burden of common risk variants for psychiatric disorders. Mol Psychiatry. 2019. 2019. https://doi.org/10.1038/s41380-019-0558-2.
19. Bouveyron C, Girard S, Schmid C. High-Dimensional Data Clustering. 2007. 2007. https://doi.org/10.1016/j.csda.2007.02.009.
20. Berg L, Bouveyron C, Girard S. HDclassif: An R Package for Model-Based Clustering and Discriminant Analysis of High-Dimensional Data. J Stat Softw. 2012;46.
21. Bertoletti M, Friel N, Rastelli R. Choosing the number of clusters in a finite mixture model using an exact Integrated Completed Likelihood criterion:1–23.
22. Chiu D, Talhouk A. DiceR: An R package for class discovery using an ensemble driven approach. BMC Bioinformatics. 2018;19.
23. Bouveyron C, Girard S, Schmid C. High-Dimensional Discriminant Analysis. Commun Stat - Theory Methods. 2007;36:2607–2623.
24. Robin X, Turck N, Hainard A, Tiberti N, Lisacek F, Sanchez J-C, et al. pROC: an open-source package for R and S+ to analyze and compare ROC curves. BMC Bioinformatics. 2011;12:77.

25. Tibshirani R. Regression Shrinkage and Selection via the Lasso. J R Stat Soc (Series B). 1996;58:267–288.
26. Friedman J, Hastie T, Tibshirani R. Regularization Paths for Generalized Linear Models via Coordinate Descent. J Stat Software, Artic. 2010;33.
27. López-Ratón M, Rodríguez-Álvarez MX, Cadarso-Suárez C, Gude-Sampedro F. OptimalCutpoints: An R Package for Selecting Optimal Cutpoints in Diagnostic Tests. J Stat Software; Vol 1, Issue 8. 2014. 13 November 2014.
28. Pollard KS, Dudoit S, van der Laan MJ. Multiple Testing Procedures: R multtest Package and Applications to Genomics, in Bioinformatics and Computational Biology Solutions Using R and Bioconductor. Springer; 2005.
29. Yee TW. The VGAM Package for Categorical Data Analysis. J Stat Software, Artic. 2010;32.
